# Exhaled particles from nanometre to millimetre and their origin in the human respiratory tract

**DOI:** 10.1101/2021.10.01.21264333

**Authors:** Gholamhossein Bagheri, Oliver Schlenczek, Laura Turco, Birte Thiede, Katja Stieger, Jana-Michelle Kosub, Mira L. Pöhlker, Christopher Pöhlker, Jan Moláček, Simone Scheithauer, Eberhard Bodenschatz

## Abstract

Detailed knowledge of the properties of exhaled particles from the human respiratory tract for all genders and ages is essential to determine the modes of transmission of airborne diseases. This applies not only to the current COVID-19 pandemic, but also to many others, be it measles, seasonal influenza or tuberculosis. To date, there are no data on the individual-specific concentrations and sizes of exhaled particles over the entire size range from nanometre to millimetre. Here we present a comprehensive data set, measured by particle size spectrometry and in-line holography covering the entire size range from 132 healthy volunteers aged 5 to 80 years for a defined set of breathing and vocalisation activities. We find age to have a large effect on small particle concentrations (<5 µm), doubling in children during adolescence and in adults over a 30-year period. In contrast, gender, body mass index, smoking or exercise habits have no discernible influence. Particles >20 µm show on average no measurable dependence on the type of vocalisation with the exception of shouting. We show evidence that particles <5 µm mainly originate in the lower respiratory tract, 5-15 µm in the larynx/pharynx, and >15 µm in the oral cavity.

---

Human exhalations contain endogenously generated particles, composed of non-volatile substances, such as salts, proteins, possibly pathogens, and water. These particles span in size from nanometre to millimetre and are referred to as aerosols and/or droplets [1–13]. Knowledge of particle size distributions for typical respiratory activities is central to understanding the transmission of airborne diseases and their control [13, 14, and references therein]. It is well known that the particle size distributions exhibit large within- and between-subject variability, as well as dependencies on respiratory activity, age and voice volume [e.g., 1, 2, 4–11, 13, 15, and references therein]. There are also considerable differences between datasets in the literature, attributable to the instruments and measurement conditions used [13]. For some particle size ranges and respiratory or vocalisation activities, the likely origin of particles in the airways has been reported, e.g. the bronchiolar fluid film rupture producing mainly submicron particles [e.g., 16, 17], production of ~(1–2) µm particles through the larynx [e.g., 15], and large particles produced by filament formation and break-up during lip opening [e.g., 18].

The scatter of absolute values of particle size distributions found in the literature, however, makes it very difficult, if not impossible, to reliably estimate the concentration of exhaled particles and thus the risk of infection from airborne diseases [e.g., see 13]. The inherent variability within and between subjects can partly explain this, but not the differences between studies. The reasons for this may be the measurement methods used and their limitations, the experimental situations, the limited number of subjects and/or also the duration of sampling. In addition, the concentration of particles with a size >20 µm still remains to be directly measured. So far, only inferred data on particle number or volume-normalised data are available [1–3, 19] or the concentration is estimated from the total exhaled particle volume [7, 20] or the exhalation flow rate [15]. In addition, there are few studies under well-controlled environmental conditions or with more than 20 subjects or with more than one breathing/vocalisation manoeuvre. Measurements of particle size distributions in children and adolescents are sparse [e.g., see 13, 21].

An assessment of the risk of infection by respiratory pathogens requires knowledge of the size distribution and concentration at the time of exhalation. The importance of the particle diameter when exhaling, *D*_0_, becomes apparent when considering that the number of pathogen copies carried by a particle scales with the particle volume, i.e.,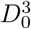 [see e.g., 14]. The shrinkage of these particles, defined as the particle diameter in the respiratory tract compared to the equilibrium size they reach at ambient relative humidity (RH), determines their deposition rates on surfaces and residence times in the air [13, 14, 22–28]. In particular, small particles, e.g. <10 µm, shrink by evaporation in a fraction of a second under typical ambient conditions and remain in the air for a long time [e.g., see 12–14, 29]. In addition, the situation-specific particle size and concentration determine the protective properties of filtering face masks [see e.g. 30, 31, and references therein] and of technical air filtration.

The exhaled particles from the oral cavity primarily consist of saliva, those from the Lower Respiratory Tract (LRT) primarily of Airway Surface Liquid (ASL). Saliva is ~ 99% water mixed with various salts and organic materials like mucins [e.g., see 13]. At a solids content of 1% and assuming that the dissolved solids have the same density as water, the shrinkage for saliva droplets is a factor of 4 to 5 at RH<40%, which is in agreement with measurements [e.g., see 13, 32]. The airways in the LRT of the adult lung has approximately 23 generations of bifurcations with the trachea being the zeroth generation and the terminal bronchioles number 23. In the LRT, the ASL has two distinct layers depending on the generation: (i) a complex hydrogel mucus layer that is directly exposed to the inhaled/exhaled air and acts as a clearance vehicle and protective barrier against foreign particles and pathogens (up to generation 15-16), and (ii) a periciliary fluid-like layer in which the cilia beat and which up to generation 15-16 is below the first layer (for generation >17 only the periciliary fluid-like layer remains) [e.g., see 33, 34]. The primary component of the overall ASL in healthy humans is water, with a nonvolatile solid fraction of approximately 1.1-2.3% *wt*[35, 36]. Using these values and assuming that the solids have the same density as water, a shrinkage factor of 3.5-4.5 can be expected for completely dried ASL. This is lower than the swell factor of 6.25 reported for exocytosed airway mucus but it is higher than the value estimated by Nicas *et al*. (2005)[23], which is based on the rather high ~ 9% *wt* solid content taken from the measurements of Effros *et al*. (2002) [37]. Holmgren et al. (2011) [5] report measurements of exhale particle size distributions measured between 5-35% and 70-85% RH and extrapolate assuming pure hygroscopic growth by approximating the ASL by an aqueous NaCl solution. They calculate a shrinkage factor of 2.4 from 99.5% to 75% RH. Recent data of Groth *et al*. (2021) [38], which are based on measurements of cough particles at RH<90% coupled with hygroscopic growth models yield a shrinkage factor of 2.8 at 0% RH. It should be noted that the data measured at high RH shown in Fig. 3 of Groth *et al*. (2021) [38] are also consistent with larger shrinkage factors. Overall, the published shrinkage factors vary greatly.

In summary, a more detailed knowledge of the concentration, size, and shrinkage of exhaled particles in all genders and age groups is indispensable to determine the modes of transmission of airborne diseases. Figure 1a summarises our measurements of particle size distributions from 50 nm to 1 mm obtained from more than 5800 minutes of aerosol spectrometry and 12000 holograms of exhaled air from 132 healthy individuals (56 female, 76 male) aged 5-80 years (see SI, Table S.1). We used particle size spectrometers (PSS) and in-line holography and measured in a better than ISO Class 4 cleanroom. Subjects performed nose/mouth breathing, normal/loud speaking, singing, humming, shouting and other specific activities (e.g. singing/shouting with open mouth). Respiratory particles larger >6 µm were directly measured with in-line holography, a proven instrument in the field of atmospheric cloud micro-physics [39], just a few centimetres from the mouth and nose of the subject; particles <10 µm were measured using the PSSs after exhalation was captured via specifically designed full-face masks or by sampling with a funnel in front of the subjects mouth/nose for the same activities (see Methods and SI, subsection 1.3 for details).The dependencies of the data on gender, age, vocal sound pressure, height and body mass index (BMI) are discussed below. Figure 1b summarises our measurements of particle size distributions for very specific activities that allow us to learn much about the origin of the particles in the respiratory tract as discussed in detail later in this manuscript and marked in Fig. 1a.

**Figure 1:**
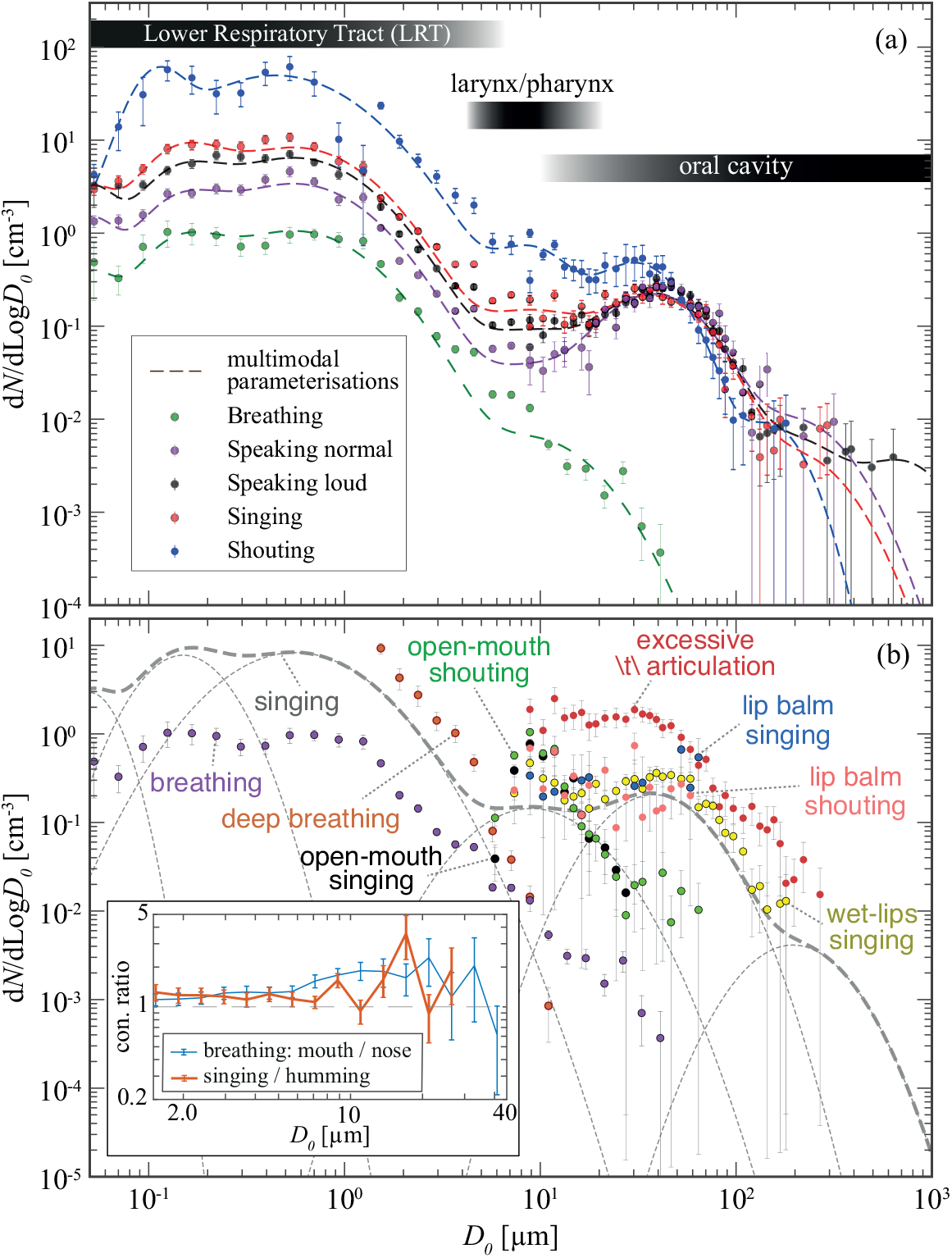
Measured exhalation particle size distributions versus exhaled particle diameter *D*_0_. **(a)** particle size distribution from the arithmetic mean of the normalised concentrations over all subjects performing the same activity. Dashed lines show multimodal lognormal fits that capture the data well for each activity (the parameters of the fits are given in Table S.4 of the SI). The lognormal functional form is based on what one would expect for the physical processes involved [e.g. 13]. The average of A-weighted-decibels dBA 3rd quartiles measured at a distance of ~ 20 cm away from the subject are 78.6 dBA, 83.6 dBA, 85.7 dBA and 102.4 dBA for speaking normally, speaking loudly, singing and shouting, respectively. Horizontal stripes above the curves indicate the inferred sites of origin in the respiratory tract. **(b)** bin-normalised particle size distributions for breathing, singing and other *special* respiratory activities (all activities are fully defined in Table S.2). The thick grey dashed line shows the multimodal parameterisation presented in Table S.4 of the SI for singing (both the data and the parameterisation are shown in a) while the thin ones visualise individual modes. The inset shows the ratio of total particle concentration between mouth-breathing and nose-breathing, and between singing and humming. The vertical bars show the (symmetrical) standard error for each diameter channel of the instrument. The multimodal fits are local representations that are closest to the data and may or may not reflect production mechanisms in the respiratory tract. For the smallest and largest particles (i.e. modes 1, 6 and 7 in Table S.4 in the SI), the fit was performed to few data points and should therefore be interpreted with care.

## Shrinkage factors for saliva and airway surface liquid

To correct our data measured in the PSSs at an RH different to that of the human respiratory tract, it was necessary to quantify the shrinkage of human saliva and ASL. To this end, we used two independent experimental approaches.

In the first approach, exhaled particles were measured during breathing and singing using PSSs with/without diffusion dryers (i.e., dry/wet) for the same group of subjects. The shrinkage factor can then be calculated by dividing the diameter of the wet particles by that of the dry particles with the same concentration, with the data shown in Fig. 2a. It should be noted that although the wet measurements were conducted by breathing directly into the inlet of the spectrometer, submicron particles evaporate inside the instrument on their way to the measurement volume thus giving a “false” shrinkage factor of about unity (as seen in Fig. 2a). For larger particle diameters above 2 µm the apparent shrinkage factor for particles produced during breathing grows to 3 and cuts off. The data does not show the plateau one would expect to observe for a “true” shrinkage factor. We attribute this to the particles during breathing being small and influenced by drying in the instrument. However, for singing the data is continued to larger particles >3 µm and plateaus around a shrinkage factor of 4.5. This value is consistent with what is expected for the solid content of saliva and ASL.

**Figure 2:**
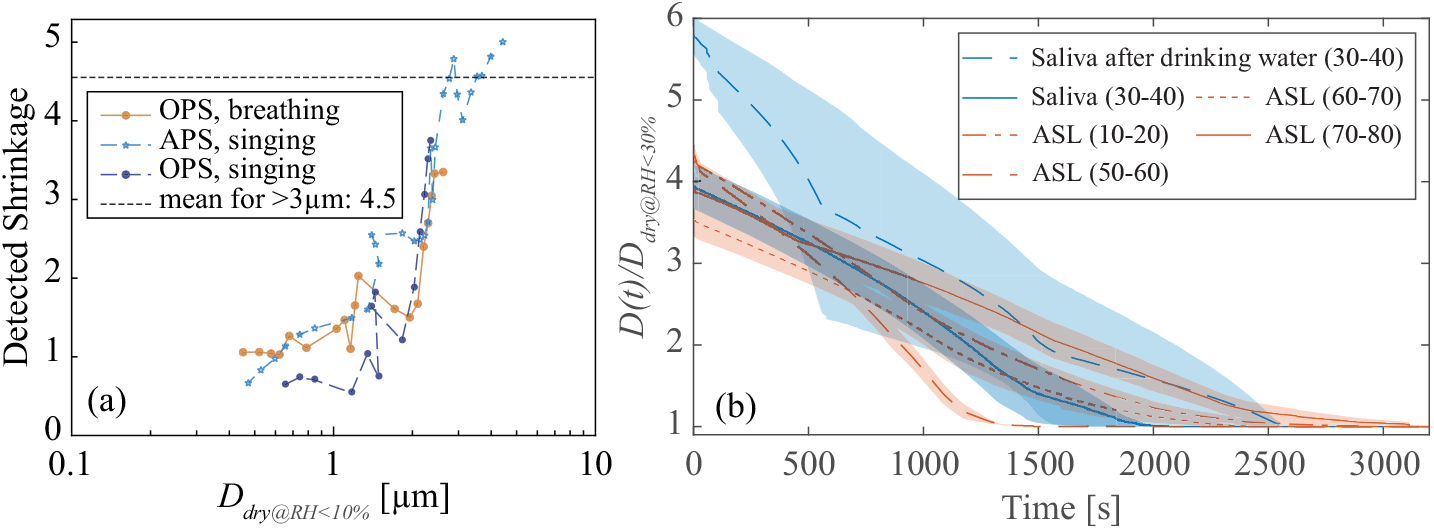
Measured Shrinkage factors measured. **(a)** By calculating the ratio between diameter of particles exhaled by a subset of subjects during breathing/singing without dryers (wet sampling) and the diameter of particles with the same concentration as the wet samples but collected with diffusion dryers (dry sampling at RH<10%); **(b)** imaging millimetre-size droplets of human saliva and ASL suspended on a human hair as they dry inside a container with RH<30% and temperature of ~ 23 °C, subject age is shown in parentheses in the legend and the shaded regions visualise the standard error. OPS stands for TSI Optical Particle Sizer model 3330 and APS stands for TSI Aerodynamic Particle Sizer model 3321. Saliva droplets were sampled either after at least 15 min without drinking water or immediately after drinking a sip of water. ASL samples were collected from four patients via tracheostomy with no respiratory-related diseases, which were examined upon collection to be free of pathogens. See Methods and subsection S.1.5 in the SI for more details.

In the second approach, we have directly measured shrinkage of human saliva and ASL as shown in Fig. 2b. The mean shrinkage factor for saliva was ~ 4, while it was ~ 5.8 when the subject drank water before sampling. This indicates that depending on the hydration level a high within-subject variability can be expected. ASL shrinkage was found to be in the range of 3.5 − 4.3. However, it should be noted that the residue-volume extraction method used here tends to underestimate the shrinkage factor, as explained in subsection S.1.5 of the SI. Putting both results together a shrinkage factor of 4.5 shown by the dashed line in Fig. 2a is consistent with both experiments and expectations based on dry mass. We use this factor in the analysis of the measured data from the PSSs.

## Respiratory particle size distribution during breathing and vocalisation

Figure 1a shows the normalised concentration of respiratory particles arithmetically averaged over all test subjects as a function of the exhaled diameter *D*_0_. The <9 µm data are obtained from the PSSs, while the data for larger particles are from the in-line holography. The shrinkage factor of 4.5 (see also Methods for details) is applied to the fully dry PSS data to calculate back the droplet exhaled diameter before merging it with the holography data. For data obtained with in-line holography, based on measurements taken a few centimetres from the subject’s mouth or nose, shrinkage is negligible. From now on, all particle diameters given refer to exhaled (wet) diameters *D*_0_, unless otherwise stated. The data from breathing and vocalisations differ significantly, i.e., vocalisation not only increased particle concentration, but also the concentration of large particles.The deviation becomes pronounced above *D*_0_ ~ 5 µm, where particle concentration for breathing decreases quickly with increasing particle size whereas for other activities it plateaus before increasing. Largest particles detected with holography across all experiments are 312 µm, 618 µm, 298 µm and 182 µm during speaking normally, speaking loudly, singing and shouting, respectively. This is below the detection limit of about 1 cm for this instrument. The concentration of <20 µm particles increases with sound pressure, which agrees qualitatively with previous observations [e.g., see 9, 10]. Looking at vocalisation at different sound pressures and single subjects rather than the mean across all subjects, the concentration of 1.5-7 µm correlates strongly with sound pressure (see SI, subsection p.2.1). This observation we attribute, at least for particles with an exhaled diameter of <5 µm, to changes in the lung volume during the inhalation and exhalation and the time gap between them at the different sound pressures rather than to the sound pressures themselves or to larynx/pharynx. The latter is discussed in detail when we address the anatomical origins of the exhaled particles in the airways. Interestingly, the particle size distribution between 20 µm-150 µm is very similar for normal/loud speaking and singing.

## Subject variability and influence of age, gender, smoking, exercising habits and Body Mass Index (BMI)

For a given activity, the variability of the PM5 concentration (particles with *D*_0_<5 µm) for a single subject follows a Gaussian distribution and can reach a factor of 10 between minimum and maximum of measured concentrations (see SI, subsection S.2.2 and Fig. S.8). It can be as high for measurements taken on the same day as for measurements taken more than 200 days apart.

For PM5, the measured variability in number concentration between subjects follows a lognormal distribution (see SI, subsection p.2.2 and Fig. p.9), which is consistent with previous observations ([e.g., see 9]). It differs between the lowest and the highest emitter by a factor of 100-150, depending on the activity. However, for 90% of subjects, PM5 concentrations are within 0.05-3.5 of the population arithmetic mean, regardless of activity. While the ratio of PM5 concentrations when breathing for the highest emitter (one of 132 subjects) to the arithmetic mean of the population is about 10, this ratio is 4.2, 4.8, 5.7 and 5.0 for normal speaking, loud speaking, singing and shouting, respectively. For all four activities - breathing, normal speech, loud speech and singing, 1.6% of the subjects studied were one standard deviation above the mean in log(*N*). The majority of subjects aged 5-14 years were among the lowest emitters in terms of PM5 concentration (5th percentile) and the majority of those aged 47-63 years were in the 95th percentile. This shows that age is an important parameter for this particle size range and vocalisation, as can also be seen in Figure 3. While it takes about 7 years for PM5 particle concentration to double in children and adolescents, this increase occurs within 30 years in adults.

**Figure 3:**
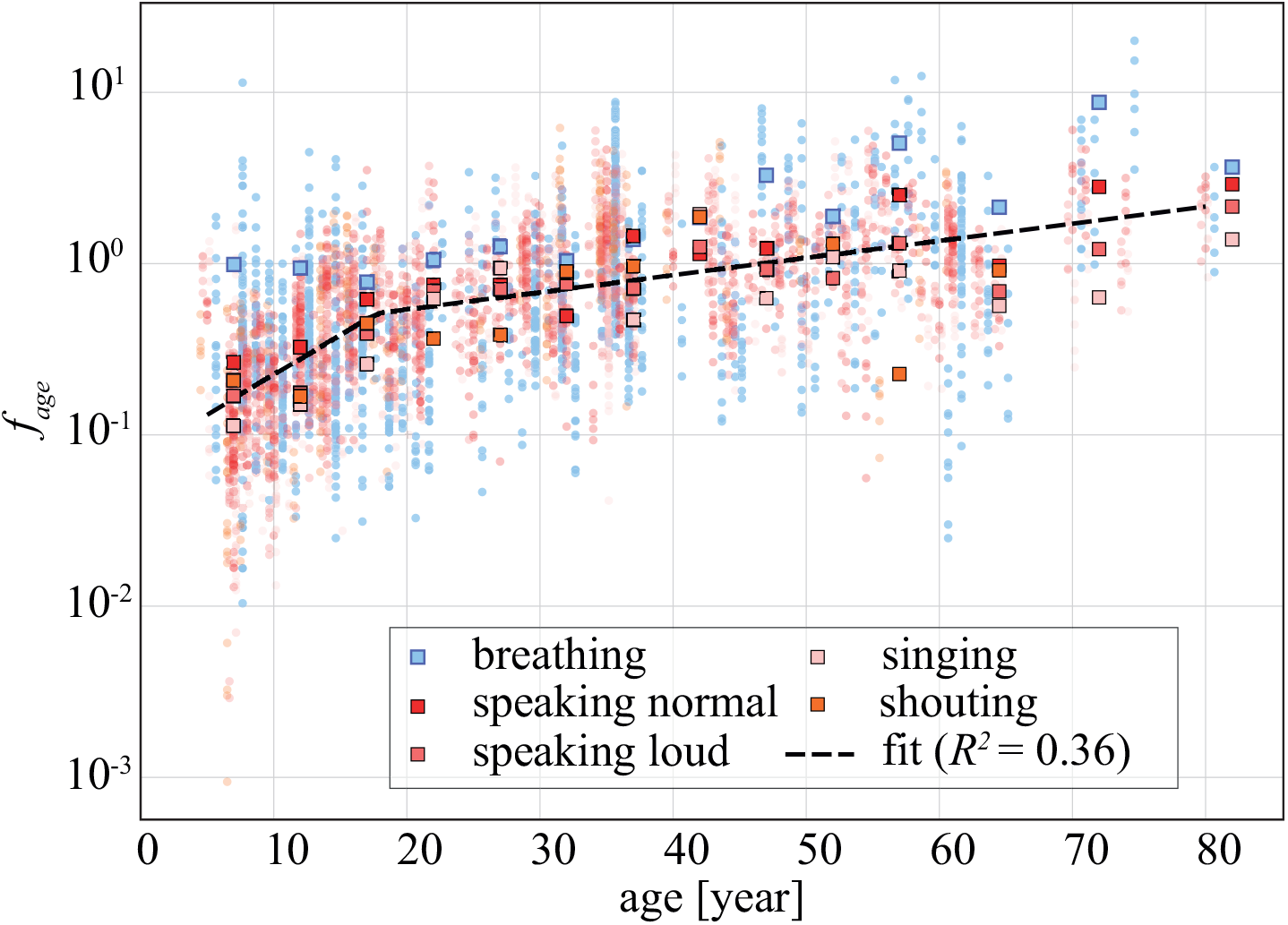
Influence of subject age on particle number concentration for different activities and particles with *D*_0_ = 1.5 µm-5.7 µm. *f*_*age*_ is defined as the number concentration produced for a given age group divided by the estimates found by the multimodal log-normal parameterisations found for the mean between all subjects shown in Fig. 1a and Table S.4 of the SI. Square symbols show the average *f*_*age*_ values in 5-year age categories for each activity, while the circle symbols, slightly shifted horizontally for each activity to be better visible, show the *f*_*age*_ for each individual experiment. The dashed line is a piece-wise linear parameterisation fitted to the the average *f*_*age*_. The fit provides a multiplier to the multimodal log-normal parameterisation to adjust for subject age: *f*_*age*_ = 10^0.047 age*−*1.12^ for younger than 18 years and *f*_*age*_ = 10^0.01 age*−*0.454^ for older subjects.

The observation that PM5 particle concentrations during vocalisation are age-dependent suggests that most PM5 particles produced during these activities are generated by the mechanism of airway closure, which is known to be age-dependent [e.g., see the detailed discussions in 8, and references therein]. In contrast, particle concentrations during normal breathing appear to be less influenced by age than during vocalisation, suggesting that in the majority of young subjects the lung volume at which airway closure occurs is not reached during normal breathing. Nevertheless, the similar shape of the concentration distribution during breathing and vocalisation suggests that the mechanisms of formation are likely to be similar. Based on the piecewise linear parameterisation shown in Fig. 3, the PM5 concentration in exhaled air of children aged 5 years during vocalisations is on average 13% of the mean determined for all subjects, while the PM5 concentration in exhaled air of an 80-year-old is 2.1 times the mean. We also find that the normalised PM5 concentration at age ~ 45 years is ~ 1, showing that our multimodal parameterisations are best at predicting PM5 concentration at this age. Depending on the activity, the correlation with age gradually disappears for particles with *D*_0_ beyond 5-8µm in both PSS and holographic data.

We found no discernible influence of gender (see SI, subsection S.2.3), smoking (see SI, subsection S.2.4) or exercise habits (see SI, subsection S.2.5) on PM5 concentration. We also could not find a conclusive dependence between BMI and PM5 concentration (see SI, subsection S.2.6).

## Origin sites within the respiratory tract

Our data from a series of carefully selected activities (see Fig. 1b) targeting a specific part of the respiratory tract show that PM5 particles originate from the LRT, whereas *D*_0_ ~5-15 µm particles originate primarily from larynx/pharynx and *D*_0_>15 µm particles primarily from oral cavity. The first point to note in Fig. 1 for PM5 particles is the strong similarity between the particle size distributions for singing and breathing. Furthermore, the correspondence in particle concentration between pure nasal and pure oral breathing (*R*^2^ > 99% and *p* < 0.01, see inset plot in Fig. 1b) renders the oral and nasal cavities as the origin of PM5 unlikely. In addition, during breathing the inactive vocal cords should not contribute significantly to particle production. As shown in Fig. 1b during deep breathing, the LRT alone is capable of producing even higher particle concentrations than those observed during singing or even coughing (see also SI subsection S.2.7 and Fig. S.16). Such a dramatic increase in particle number concentration during deep breathing is likely triggered by the subject reaching the point at which extensive airway closure occurs, which has also been reported in several other previous studies [see the detailed history on this topic in 8]. It is also known that airway closure occurs at shallower lung volumes with age [8]. We have also found that the breathing frequency does not influence the exhale particle size distribution (see SI, subsection S.2.9.3). Furthermore, pausing between full inhalation and exhalation significantly decreases particle emission similarly to observations reported previously [e.g., 4, 5] (see SI, subsection S.2.9.4). We conclude from these observations that PM5 particles during breathing originate predominantly from the LRT, i.e., the lung and trachea, and their concentration is not a function of breathing frequency but the lung volume and the pause between inhalation and exhalation.

Now the question arises, where in the respiratory tract do the PM5 particles originate from during singing (and other vocalisations)? Surely some of these particles are produced in the LRT similarly to breathing, since the subjects not only vocalised but also had to breathe. Thus, also during singing and shouting the LRT must be a major PM5 contributor. But what is the influence of the remaining respiratory tract? The similarity in particle size distribution between humming the “Happy Birthday” song with mouth closed and singing the same song shown in the inset of Fig. 1b suggests that PM5 are not produced in the oral cavity during singing. This leaves the larynx and pharynx as the remaining candidates next to the LRT. Although there is a weak correlation between PM5 concentration and subject sound pressure (see SI, subsection S.2.1, Figure S.7), which would support the assumption that the larynx is a PM5 source, the large scatter in the data observed suggests strongly that the larynx/pharynx is not a major source of PM5 production. In addition, it is rather unlikely that acoustic waves generated during vocalisations could stimulate extra particle production in the LRT (see SI, subsection S.2.9.5). We also found no significant difference in particle size distribution between breathing with and without vocalisations at fixed exhalation flow rate and lung capacity (see SI, subsection S.2.9.6 for a more detailed discussion), and most importantly we have found that PM5 consistently arrive 1-2 s later than the larger particles regardless of activity. This time delay strongly suggests that PM5 are produced mostly within the LRT (see SI, subsection S.2.9.7). Finally, the dependency between PM5 concentration and subject age is consistent with the LRT being the main origin site for PM5. From these observations, we conclude that PM5 are predominantly produced in the LRT for all of the activities studied here.

Particles >5 µm are mostly produced during vocalisation, so the likely sites of origin are the pharynx/larynx, nasal cavity and oral cavity. This is supported by the fact that we could not detect any >6 µm particles (i.e., the lower detection limit of holographic setup) while measuring various nose/mouth breathing and humming manoeuvres. When singing and shouting with the mouth open, which should eliminate lip contributions, we found that the larynx/pharynx (and possibly the tongue) are very effective in producing ~5-15 µm particles. The higher particle concentration observed when singing with the mouth open compared to normal singing even suggests that some of the particles produced by the larynx/pharynx are obstructed by the oral cavity before exhalation. This obstruction could also explain part of the increase in concentration when shouting compared to singing, as the mouth is usually held open longer when shouting.

The sharp concentration drop at 15 µm for singing and shouting with the mouth open indicates that the majority of >15 µm particles detected for standard vocalisations are produced by the tongue, tongue-teeth interactions and lips. An example of particle size distribution produced only by the tongue-teeth can be seen in open-mouth sound \t\ articulation that leads to production of a wide range of particles mostly >10 µm. Singing while frequently wetting lips with the tongue lead to a particle size distribution similar to singing normally with slightly higher concentration for >10 µm particles, suggesting particles produced by the lips span a wide range of sizes too. However, the main contribution from lips becomes evident for the singing and shouting experiments when subjects applied lip balm to their lips. Applying lip balm has been shown to temporarily hinder formation of saliva filaments between the lips and reduce particle emission [18]. Figure 1b shows that applying lip balm is effective mostly for reducing emission of >75 µm particles. Taking all these observations together, the most important points of origin can be derived as a function of particle size, shown as horizontal dark stripes at the top of the figure 1a.

## Summary

With this knowledge of the size-dependent origin of particles in the respiratory tract, biological sampling of the aerosols exhaled by a person, which can be easily performed with a suitable face mask and a commercially available cascade impactor, can provide insights into the health status of different regions of the respiratory tract. With the comprehensive reference data set presented here on exhaled particles for all genders and age groups, it is also possible to better determine the modes of transmission of airborne diseases, assess the risk of infection and develop disease control strategies. This is important not only for the current COVID-19 pandemic, but also for many others, be it measles, seasonal flu, tuberculosis, or infectious diseases that are yet to come.

## Methods

### Subjects and activities

A total of 132 healthy volunteers aged between 5 and 80 years were tested. They all participated voluntarily, were informed in advance about the conduct of the experiment and subsequently consented to their participation. The subjects had to be legally competent and not impaired to carry out activities in this study. For children, both the children themselves and their legal guardians gave consent. Participation could be revoked at any time during the study. The study was approved by the ethics committee of the Max Planck Society. The subjects were recruited in various ways: via the homepages of the Max Planck Institute for Dynamics and Self-Organisation (MPIDS), the Institute für Hospital Hygiene and Infectiology at the University Medical Center Göttingen Georg-August-Universität (UMG), via the investigators themselves and there were also active requests with the wish to participate that could be considered. The data were pseudonymised in accordance with the approved data protection concept and anonymised from the data extraction step onwards. An overview of the age distribution of the subjects and a description of the activities performed can be found in the SI, section S.1.1 and S.1.2. The subjects performed different breathing activities and the corresponding particle size distributions were measured with different particle characterisation instruments (see SI, table S.1).

### Cleanroom

All measurements were performed in a nominal ISO Class 6 cleanroom (i.e. less than 1 million particles per cubic metre). However, our measurements show that the cleanroom met at least ISO Class 4 criteria, i.e. less than 1000 >0.3 µm particles per m^3^. The average air temperature and RH in the cleanroom were about 22 °C and 45%, respectively. The cleanroom was separated from the outside air by an airlock. In this airlock, the cleanroom clothing (cleanroom gown with bonnet, powder-free sterile gloves, boots and FFP2 face mask) was put on before entering the “isolated side” of the cleanroom and the devices and equipment brought into the cleanroom were thoroughly cleaned here. In the cleanroom powder free paper was used for recording the measurements. The cleanroom air was constantly monitored during the tests to ensure that the background air was ISO Class 4 conditions. As a rule, only one test person (plus one accompanying person for children and adolescents) and a maximum of three scientists stayed in the room during the experiments. All persons in the cleanroom except for the test subject wore an FFP2 mask during the measurements.

### Instrumentation

Dried particles (as described later) with diameters ranging from 0.01 to 0.42 micrometres (in 13 log-equidistant bins) were measured with a NanoScan Scanning Mobility Particle Sizer (SMPS, model 3910, TSI Inc. Shoreview, MN, USA), while those with optical diameters (based on Mie’s spherical scattering profiles) between 0.3 µm and 10 µm (in 16 log-equidistant bins) were measured with an Optical Particle Sizer (OPS, model 3330, TSI Inc.). A sampling interval of 60 s was chosen for the PSSs. Due to problems with the SMPS instruments, not all SMPS data from all subjects can be used, whereas the OPS data are available for all subjects. Therefore, values for the particle size distribution of <300 µm are missing in our database for many children and adolescents.

In addition, an Aerodynamic Particle Sizer (APS, model 3321, TSI Inc.), 0.5-20 µm) was used for a large part of the experiments to measure the particle size distribution simultaneously with the OPS (and SMPS). We found close agreement between the optical diameters derived from the OPS and the aerodynamic diameter of the APS for <4.0 µm particles. However, the APS detection efficiency was very low for larger particles, which was also reported in previous studies [e.g. see 13, and references therein]. For this reason, we did not use the APS data, as we had verified with the APS that the OPS data were accurate. We also matched the OPS data with a GRIMM aerosol spectrometer model 11-D using dolomite dust, glycerol mist with 0.5% NaCl and respiratory particles, and the normalised concentrations were within 0.5 to 1.5Ȧll instruments used here had valid factory calibrations.

We also compared the concentration values from the last bin of the SMPS with the first bin of the OPS, whose particle size range overlaps. It was found that the concentrations measured by the SMPS are higher than those of the OPS. This discrepancy was expected since, according to calibration certificates, the detection efficiency of our OPS unit for the smallest bin is 50%, while the largest bin of our SMPS has an efficiency of 90%. After applying the associated detection efficiency, the value of the merged channel was within ~ 14% of the values measured in the last chamber of the SMPS. Finally, based on factory and internal calibrations, we approximated the geometric diameter of the particles with the optical diameters of the OPS and the SMPS electron mobility diameters with an average deviation of <15%.

The particle size distributions of large particles were measured using HALO-Holo, which is a particle imaging sensor using in-line holography with an effective pixel size of 2.96 µm and a 160 mm distance between the two arms [39]. Images were taken with a 6576 × 4384 pixel CCD camera with a frame rate of 6 frames per second. The volume sampling rate was 230 cm^3^ s^*−*1^. After numerical reconstruction [see 40, for more detail] and classification of objects via supervised machine learning, the size, shape and location of particles between 6 µm and a few millimetres could be determined. Under the laboratory conditions used here, the image background is more stable and small particles are easier to detect than those obtained during airborne measurements of atmospheric clouds. We have also performed a size calibration with NIST-traceable glass microspheres from 8 µm to 50 µm particle diameter (see SI, subsection S.1.7) and found a maximum sizing uncertainty of ± 1.0 µm. Concentration measurements were previously found to be in agreement with optical particle spectrometers for ± >10 µm particles [39]. We have also calculated the HALOHolo detection efficiency relative to near-camera regions. It was found that in a region 2.5-20 mm away from the central plane and 2.5 mm away from the probing volume edges, relative detection efficiency is 87% for 6 µm, >90% for >12 µm, and nearly 100% for >32 µm particles. However, as a compromise between statistical convergence, which requires large sampling volume, and uniform detection efficiency for all particle size, which is achieved for a small portion of probing volume, we have restricted our analyses to a 60 mm long region in the centre of sampling volume. With this the “effective sampling volume” is 14.5 mm wide by 9.6 mm high by 60.0 mm long, which at 6 Hz sampling frequency amounts to sampling rate of 3 l min^*−*1^. In order to calculate representative concentration values for each respiratory activity, we accumulated data from holograms that had at least one particle in the effective sampling volume and then divided the total particle count in each size bin by the product of the volume and number of holograms contributed to the accumulated particle count.

Empty holograms were excluded in order not to count situations in which the subject did not speak/exhale into the sampled volume. Due to the extreme directionality of respiratory flow, it was non-trivial for the subjects to always target the holographic sampling volume throughout the duration of the measurements, even though the subjects were monitored throughout the measurement. As the holograms can only be analysed in post-processing we did not have a direct control at the time of the experiment. As a result the empty holograms were designated as *false zero* holograms, the inclusion of which would have affected the calculated concentration by a factor of 4 to 40 (depending on the activity) lower than those shown in Fig. 1a. The inclusion of holograms with at least one particle within the total probe volume (as opposed to including holograms with at least one particle in the effective sampling volume) would have affected the concentration of (>9 µm) particles in Fig. 1a reduced by a factor of ~ 2.3, which in any case is within the variability between subjects. Nevertheless, the observed agreement between the measured concentration of particles with 9 µm exhaled diameter with the PSS and HALOHolo supports confidence in the correctness of the analyses and the validation procedure used in HALOHolo.

For sound pressure measurements we used the 2 Hz PEAKTECH 8005 digital sound-level meter capable of measuring sound levels between 0.1 dB-130 dB at 0.1 dB resolution. Only the sound pressure data that were obtained during the combined holographic and funnel-sampling measurements were used, during which unobstructed sound pressure at a distance of about 20 cm away from the subject was measured. Nonetheless, absolute values reported here should be taken cautiously since the walls of the cleanroom were acoustically reflective.

### Measurement setup

While for holographic measurements respiratory particles were directly imaged at about <5 cm away from the subject’s mouth, for PSS measurements the exhale flow was sampled via two different methods, namely (i) a specifically designed full-face mask, hereafter referred to as “isolation shield”, and (ii) plastic funnel (see SI, Figure S.1). The (i) isolation shields were modified snorkel masks and had different flow paths for in- and exhalation that were controlled by oneway valves. With an in-house designed adaptor, we sampled the exhaled air directly (see SI, subsection S.1.3). Two different isolation shield models were used, a comparison in measured particle size distribution is shown in the SI, Fig. S.19. The (ii) funnel has a diameter of 15 cm and was held approximately 10 cm in front of the subjects face at the height of mouth and nose (holographic probing volume simultaneously in between).

Sampling tubes connecting the funnel/isolation-shield to the PSSs were all electrically conductive (anti-static) PTFE tubing conforming with EN 12115. The sampled air from the funnel/isolation-shield went first through two Grimm diffusion dryers model 8913 in series (each 29 cm long with 19 cm outer diameter), then to a flow-splitter and finally to the PSSs (see SI, Fig. S.1) The Grimm dryers were filled with silica beads to dry sampled air to *RH* below 30% (i.e., below the efflorescence RH, see [13]) to ensure measured particles are fully dried. The silica beads were replaced with new ones from time to time. Without any diffusion dryer, it would take approximately 4 min until the air inside the tubing is close to water saturation (see SI, Figure S.5). Even one diffusion dryer is sufficient to reduce *RH* to values below 20%, with two diffusion dryers the equilibrium *RH* even during a breathing, speaking or singing experiment is approximately 10%.

### Data corrections

Due to the addition of diffusion dryers on the sampling tubes, it is expected that some particles are lost before reaching the PSSs. To compensate these losses and correct the data, we have performed a series of controlled experiments in which an OPS measured particle concentration in a well-mixed room filled with dolomite dust or 0.5% glycerol through diffusion dryers (and tubing), while at the same time another OPS was measuring particle concentration in the room through a tube of similar total length and curvature. The OPSs were first placed in the well-mixed room very close to each other without any tubing to cross-check and correct their measurements against each other in order to ensure they produce similar results once they are in similar conditions. It was found that particle loss in the driers was almost independent of particles size and is about 30% for dried glycerol/NaCl particles and 19% for the dolomite dust, i.e., an average loss of ~ 24%. To correct the cleanroom data for particle loss in the dryers we multiplied all PSS concentration values by a factor of 1.24.

We have also found that the concentration of dry sub-micron particles, i.e., *D*_0_ < 5 µm, in the samples measured with the isolation shield is on average about 2.6 times higher than in the samples collected via the funnel for the same subject/activity combinations. This we associate with the fact that with the isolation shield a lower sample dilution with the cleanroom air can be expected than with the funnel, which is achieved by using a *buffer volume* in sampling line of the isolation shield (more detail is presented in subsection S.2.9.2 in the SI). We also found that the concentration measured with the isolation shield and the dryers, on one hand, and measured by directly exhaling into the OPS (and without dryers), on the other, are close to unity (~ 1.2 − 1.4) for fully-dried <1 µm particles, indicating that possible electrostatic losses due to the plastic components of the isolation shields are negligible. By using the Particle Loss Calculator (PLC) tool [41] while taking into account all possible loss mechanisms and using conservative values for inlet aspiration angles and flow rate, and sampling-tube length and angle of curvature/incidence to assess the worst-case sampling efficiency, we found that the total sampling efficiency for <2 µm particles is >70%, which is an acceptable value. For larger particles PLC estimates a sharp decrease in sampling efficiency due to inertial impaction. This is, in particular, noticeable when comparing the concentration of dry particles larger than 2 µm between the samples collected by the funnel and those collected by the isolation shield, for which strong inertial loss on the frontal part of the isolation shield is expected, i.e., the ratio of particle concentration measured by the isolation shield to that of the funnel is about 1.0 for 2 µm, 0.4 for 3 µm and 0.05 for 10 µm dry particles. Considering all the above mentioned points, it is evident that our PSS data does not capture the true concentration for >~ 2 µm dry particles (>~ 9 µm exhaled diameter), hence, the final data shown in Fig. 1a are obtained by replacing >~ 2 µm dry particles with those obtained by the holographic setup with fully wet particles. Other advantages and disadvantages of different sampling methods (e.g., whether or not isokinetic sampling is achieved for different size ranges) are discussed in detail in SI, subsection S.1.3.

Furthermore, given the model presented in 3 and the non-uniform age distribution shown in Table S.1 for the standard activities between SMPS (average median age of ~ 35) and OPS (average median age of ~ 26), estimations of the multimodal parameterisation for PM5 particles is associated with ~ 20% uncertainty due to non-uniformity in the age distribution. However, the data is not corrected due to this since the variability within and between subjects is much larger than this uncertainty.

### Measurement procedure

Subjects were investigated one at a time. After wearing the full cleanroom suits, hood and shoes in the cleanroom airlock and entering the isolated side of the cleanroom, first the isolation shield was adjusted to fit the subject’s face properly. We had isolation shields in two different sizes and took the size that was matching the subject’s face best. For a few subjects the seal around the face was not perfect, hence, the leaky parts were filled with lint-free cloth until leak-tight. The tubing, filters and dryers were all replaced with new ones for each subject to minimise the risk of infection and contamination of measurements between subjects. The subject typically started with isolation-shield measurements and then went on with simultaneous funnel and holographic measurements. Until the subjects were ready for the first measurements they had already spent a few minutes (~ 5-10 minutes) inside the cleanroom. As a result, their lung should have been cleared of non-respiratory-origin particles they could have inhaled in the outside air. Standard activities carried out in sequence with isolation shield were breathing through the nose, breathing through the mouth, reading a phonetic-standard text with normal and loud voice, singing “happy birthday to you” with arbitrary names and (for some subjects) humming “happy birthday to you” (see SI, subsection 1.2 for more details). Each activity was carried out for 3-5 min at least for isolation shield measurements. Some subjects were also willing to perform shouting “goal” or its German equivalent “tor” or various other manoeuvres and forced-coughing activities, each for 1 min. The funnel/holographic experiments included reading and singing for about two minutes (shouting and coughing one minute each) excluding breathing-related activities. In between activities subjects were allowed to take a break or drink water at will.

## Data Availability

Data is available upon reasonable request and after acceptance of open data repositories and on aerosol.ds.mpg.de

https://aerosol.ds.mpg.de

## Acknowledgements

The authors would like to thank Udo Schminke and the machine shop of the Max Planck Institute for Dynamics and Self-Organisation (MPIDS) for their technical support during the campaign. We would also like to thank Freja Nordsiek from the MPIDS for fruitful discussions and analysing HALOHolo detection efficiency, Katja Walter and Sigrid Clauberg from the University Medical Center Göttingen (UMG) for assisting with the measurements on human subjects, Sarah Romanowski from the MPIDS for maintaining the cleanroom and providing the cleanroom equipment, Katharina Gunkel and Andreas Renner from the MPIDS for maintaining and disinfecting the isolation shields, Artur Kubitzek from the MPIDS for technical support and maintaining equipment, Jan-David Förster from them Max Planck Institute for Chemistry for participating in the campaign in February and Andreas Beste from the UMG for providing the ASL samples. We would like to thank our volunteers who came from the MPIDS, the UMG, the Hannover State Opera, the Hanonver University of Music Theatre and Media, the Jacobi Kantorei Göttingen, the Brass Band Association of North Rhine-Westphalia, the Musikverein Oesdorf e.V., the Landeskoordinator Klassenmusizieren, the Evangelischer Posaunendienst and the local schools in Göttingen and Bad Sachsa. We would also like to thank the Max Planck Society for making research in this field possible.

## Author Contributions

GB, EB, SS, OS, BT, LT designed the experimental procedures for the subject measurements. JM designed and conducted the ASL and saliva measurements. SS advised all medical aspects of the investigation. EB wrote the ethics application. MP, CP consulted on the experiments. GB, EB, JK, OS, KS, BT, LT conducted the experiments on the human subjects. GB, OS, BT with the help of EB analysed and interpreted the data, OS and GB prepared the holography data. SS, MP, CP consulted on the interpretation of the data. GB with EB wrote the draft of the main text and GB, OS, BT with EB wrote the draft of the supplementary information. All authors participated in writing the final text.

## Data availability

The datasets generated and/or analysed during the current study are available from the corresponding authors on reasonable request. A concise version of the dataset is freely available in the HEADS (Human Emission of Aerosol and Droplet Statistics) web-app at https://aerosol.ds.mpg.de/en/.

## Ethics statement

The non-invasive exhaled aerosol sampling study which led to this paper was approved by the Ethics Commission of the Max Planck Society (Submission 2020 23). All subjects gave their written consent to data storage and analysis.

## References

[1] Duguid, J. P. The size and the duration of air-carriage of respiratory droplets and droplet-nuclei. Epidemiol. Infect. 44, 471–479 (1946).

[2] Loudon, R. G. & Roberts, R. M. Droplet expulsion from the respiratory tract. Am. Rev. Respir. Dis. 95, 435–442 (1967).

[3] Xie, X., Li, Y., Sun, H. & Liu, L. Exhaled droplets due to talking and coughing. J. Roy. Soc. Interface 6, S703–S714 (2009).

[4] Johnson, G. R. & Morawska, L. The mechanism of breath aerosol formation. J. Aerosol Med. Pulm. D. 22, 229–237 (2009).

[5] Holmgren, H., Bake, B., Olin, A.-C. & Ljungström, E. Relation between humidity and size of exhaled particles. J. Aerosol Med. Pulm. D. 24, 253–260 (2011).

[6] Morawska, L. et al. Size distribution and sites of origin of droplets expelled from the human respiratory tract during expiratory activities. J. Aerosol Sci. 40, 256–269 (2009).

[7] Chao, C. et al. Characterization of expiration air jets and droplet size distributions immediately at the mouth opening. J. Aerosol Sci. 40, 122–133 (2009).

[8] Bake, B., Larsson, P., Ljungkvist, G., Ljungström, E. & Olin, A.-C. Exhaled particles and small airways. Respir. Res. 20, 8 (2019).

[9] Asadi, S. et al. Aerosol emission and superemission during human speech increase with voice loudness. Scientific Reports 9, 2348 (2019).

[10] Alsved, M. et al. Exhaled respiratory particles during singing and talking. Aerosol Sci. Tech. 54, 1245–1248 (2020).

[11] Merghani, K. M. M., Sagot, B., Gehin, E., Da, G. & Motzkus, C. A review on the applied techniques of exhaled airflow and droplets characterization. Indoor Air 31, 7–25 (2021).

[12] Randall, K., Ewing, E. T., Marr, L., Jimenez, J. & Bourouiba, L. How Did We Get Here: What Are Droplets and Aerosols and How Far Do They Go? A Historical Perspective on the Transmission of Respiratory Infectious Diseases. SSRN (2021).

[13] Pöhlker, M. L. et al. Respiratory aerosols and droplets in the transmission of infectious diseases. 2103.01188 (2021).

[14] Nordsiek, F., Bodenschatz, E. & Bagheri, G. Risk assessment for airborne disease transmission by poly-pathogen aerosols. Plos One 16, 1–41 (2021).

[15] Johnson, G. et al. Modality of human expired aerosol size distributions. J. Aerosol Sci. 42, 839–851 (2011).

[16] Johnson, G. R. & Morawska, L. The mechanism of breath aerosol formation. J. Aerosol Med. Pulm. D. 22, 229–237 (2009).

[17] Almstrand, A.-C. et al. Effect of airway opening on production of exhaled particles. J. Appl. Physiol. 108, 584–588 (2010).

[18] Abkarian, M. & Stone, H. A. Stretching and break-up of saliva filaments during speech: A route for pathogen aerosolization and its potential mitigation. Phys. Rev. Fluids 5 (2020).

[19] Han, Z., Weng, W. & Huang, Q. Characterizations of particle size distribution of the droplets exhaled by sneeze. J. Roy. Soc. Interface 10, 20130560 (2013).

[20] Smith, S. H. et al. Aerosol persistence in relation to possible transmission of sars-cov-2. Phys. Fluids 32, 107108 (2020).

[21] Muürbe, D. et al. Aerosol emission of adolescents voices during speaking, singing and shouting. Plos One 16, e0246819 (2021).

[22] Wells, W. F. et al. On air-borne infection. study ii. droplets and droplet nuclei. Am. J. Hyg. 20, 611–18 (1934).

[23] Nicas, M., Nazaroff, W. W. & Hubbard, A. Toward Understanding the Risk of Secondary Airborne Infection: Emission of Respirable Pathogens. J. Occup. Environ. Hyg. 2, 143–154 (2005).

[24] Xie, X., Li, Y., Chwang, A., Ho, P. & Seto, W. How far droplets can move in indoor environments–revisiting the wells evaporation-falling curve. Indoor Air 17, 211–225 (2007).

[25] Bourouiba, L., Dehandschoewercker, E. & Bush, J. W. Violent expiratory events: on coughing and sneezing. J. Fluid Mech. 745, 537–563 (2014).

[26] Miller, S. L. et al. Transmission of SARS-CoV-2 by inhalation of respiratory aerosol in the Skagit Valley Chorale superspreading event. Indoor Air 31, 314–323 (2021).

[27] Chong, K. L. et al. Extended lifetime of respiratory droplets in a turbulent vapor puff and its implications on airborne disease transmission. Phys. Rev. Lett. 126, 034502 (2021).

[28] Wang, J. et al. Short-range exposure to airborne virus transmission and current guidelines. PNAS 118 (2021).

[29] Wang, C. C. et al. Airborne transmission of respiratory viruses. Science 373, eabd9149 (2021).

[30] Cheng, Y. et al. Face masks effectively limit the probability of SARS-CoV-2 transmission. Science (2021).

[31] Bagheri, G., Hejazi, B., Thiede, B., Schlenczek, O. & Bodenschatz, E. Face-masks save us from SARS-CoV-2 transmission. 2106.00375 (2021).

[32] Lieber, C., Melekidis, S., Koch, R. & Bauer, H.-J. Insights into the evaporation characteristics of saliva droplets and aerosols: Levitation experiments and numerical modeling. J. Aerosol Sci. 154, 105760 (2021).

[33] Song, D., Cahn, D. & Duncan, G. A. Mucin biopolymers and their barrier function at airway surfaces. Langmuir 36, 12773–12783 (2020).

[34] Romanò, F., Muradoglu, M., Fujioka, H. & Grotberg, J. The effect of viscoelasticity in an airway closure model. J. Fluid Mech. 913, A31 (2021).

[35] Hill, D. B. et al. A biophysical basis for mucus solids concentration as a candidate biomarker for airways disease. Plos One 9, e87681 (2014).

[36] Anderson, W. H. et al. The relationship of mucus concentration (hydration) to mucus osmotic pressure and transport in chronic bronchitis. Am. J. Resp. Crit. Care 192, 182–190 (2015).

[37] Effros, R. M. et al. Dilution of respiratory solutes in exhaled condensates. Am. J. Resp. Crit. Care 165, 663–669 (2002).

[38] Groth, R., Cravigan, L. T., Niazi, S., Ristovski, Z. & Johnson, G. R. In situ measurements of human cough aerosol hygroscopicity. J. Roy. Soc. Interface 18, 20210209 (2021).

[39] Schlenczek, O. Airborne and ground-based holographic measurement of hydrometeors in liquid-phase, mixed-phase and ice clouds. Ph.D. thesis, Particle Chemistry, Max Planck Institute for Chemistry, Max Planck Society (2018).

[40] Fugal, J. P., Schulz, T. J. & Shaw, R. A. Practical methods for automated reconstruction and characterization of particles in digital in-line holograms. Meas. Sci. Technol. 20, 075501 (2009).

[41] von der Weiden, S.-L., Drewnick, F. & Borrmann, S. Particle loss calculator – a new software tool for the assessment of the performance of aerosol inlet systems. Atmos. Meas. Tech. 2, 479–494 (2009).

