## Supplementary material for "Exhaled particles from nanometre to millimetre and their origin in the human respiratory tract": SI

#### S.1 Supplementary Methods

##### S.1.1 Subjects

We have characterised respiratory particles in exhale of 132 subjects aged between 5 and 80 years with 21 subjects (12 female) 5-9 years old, 24 subjects (10 female) 10-14 years old, 16 subjects (8 female) 15-19 years old, 19 subjects (10 female) 20-29 years old, 16 subjects (5 female) 30-39 years old, 13 subjects (6 female) 40-49 years old, 12 subjects (3 female) 50-59 years old, 7 subjects (0 female) 60-69 years old, 4 subjects (1 female) 70-79 years old, 1 subjects (0 female) 80-89 years old. Subjects performed different activities with different combinations of instruments, thus, the subject/activity/instrumentation distribution is non-uniform as shown in Table S.1. The Table also shows the total sampling duration for each activity/instrumentation contribution.

| Activity | No. Subj. | Age Min-Max (Med.) | No. Samp. | No. Exps. (female%) |
|---|---|---|---|---|
| **SMPS and APS** | | | | |
| breathing | 40 | 21-64 (35) | 218 | 47 (36) |
| norm. speak | 41 | 21-64 (35) | 285 | 87 (40) |
| loud. speak | 41 | 21-64 (35) | 281 | 87 (39) |
| singing | 41 | 21-64 (35) | 247 | 89 (44) |
| shouting | 6 | 31-64 (38) | 10 | 10 (20) |
| **OPS** | | | | |
| breathing | 131 | 5-80 (27) | 1050 | 234 (37) |
| norm. speak | 129 | 5-80 (22) | 1005 | 251 (42) |
| loud. speak | 132 | 5-80 (24) | 1035 | 257 (41) |
| singing | 132 | 5-80 (28) | 809 | 281 (40) |
| humming | 60 | 5-70 (14) | 207 | 66 (42) |
| shouting | 40 | 5-61 (31) | 56 | 56 (28) |
| **Holography** | | | | |
| norm. speak | 88 | 5-80 (28) | 897 | 91 (37) |
| loud. speak | 91 | 5-74 (30) | 2162 | 98 (40) |
| singing | 81 | 5-74 (35) | 1700 | 95 (40) |
| shouting | 4 | 35-61 (36) | 309 | 9 (0) |

Table S.1: Overview of subjects and samples analysed per activities and instrument. Number of samples for measurements carried out with PSSs is the total duration of measurements in minutes while for the those carried out with in-line holography is the total number of holograms with particles in the central region (i.e., the effective sampling volume).

##### S.1.2 Respiratory activities investigated

A summary of all respiratory activities discussed in this manuscript is given in Table S.2. Each of them was performed for one minute at minimum. The times for inhalation or between words/phrases in shouting or between coughs were not documented but can be retrieved from the time series data of the sound pressure meter if needed. For the speaking activities, different texts were used, depending on the subject's age and reading abilities. Adults read either the English standard text "Arthur the Rat" (from `https://www.york.ac.uk/media/languageandlinguistics/documents/currentstudents/linguisticsresources/Standardised-reading.pdf`) or the German standard text "Der Nordwind und die Sonne" plus "Unser Garten" (from Bergauer/Janknecht: Praxis der Stimmtherapie, 3rd Ed.). The youngest subjects were reading the first three paragraphs of the German fairy tale "Die Bremer Stadtmusikanten" (from `https://www.familie.de/kleinkind/maerchen/die-bremer-stadtmusikanten-grimms-maerchen`).

For singing the subjects were asked to sing "Happy Birthday to You" with names chosen by the subjects themselves, in the key and pitch the subjects felt most comfortable with. Some subjects also selected individual pieces of music or songs for singing after the "Happy Birthday", which we compared against singing "Happy Birthday". For some subjects, we found a slightly higher emitted number concentration for the chosen song in comparison to "Happy Birthday", but the shape of the size distribution was the same.

Besides normal breathing, some other breathing patterns were investigated on fewer subjects. A common set of breathing patterns done by most subjects was breathing solely through the nose for some minutes, then breathing solely through the mouth. No nose clip was used for the experiment "breathing through the mouth", we relied on the subjects.

For a set of other experiments, the breathing pattern was synchronised with a "breathing visualisation video" showing the deep inhalation up to total lung capacity (TLC) and deep exhalation down to lung's residual volume (RV) graphically (from `https://www.youtube.com/watch?v=aXItOY0sLRY`). This was done at normal speed, double speed, half speed and quarter speed to also investigate the role of the breathing frequency and the total volume flux.

##### S.1.3 Measurement Setups

In Figure S.1, the different methods of how the exhaled air is sampled for analysis with the PSSs are shown. In the simplest setup **c**, a funnel with 15 cm diameter was held approximately 10 cm in front of the subjects face at the height of the mouth and nose. The funnel was used in combination with the holographic setup (described in main paper), whose measurement volume was placed in between subject and funnel (Fig. S.3**c**). Throughout the whole measurement campaign, different setups (mouthpiece/ wire) were used to make sure the distance between subject and holographic measurement volume was kept constant at 5 cm. To sample the exhaled air directly and reduce mixing with cleanroom air, full-face snorkel masks, referred to as isolation shields throughout this paper, were equipped with newly designed adaptors. Two different models were used, they are called "old" (Neuluft Panorama Snorkel Mask) and "new" (Khroom Sports Seaview X M-1502) isolation shields in this paper and are shown in Figure S.2 and S.3**a** and **b**. The isolation shields are sealed to the rim of the face and

| Activity | Description |
|---|---|
| breathing | Normal tidal breathing under relaxed conditions (nose, mouth or both), depth and rate chosen by the subject. Pure nose or mouth breathing is indicated in the data set |
| speaking normal | Reading text aloud in a sound volume comparable to conversation in quiet environment |
| speaking loud | Reading text aloud in a sound volume comparable to theatre performance on stage |
| singing | Singing “Happy Birthday” very loudly (fortissimo), tempo, key and pitch chosen by subject |
| shouting | Shouting various things (maximum loudness) like “Tor”, “Goal” or entire phrases |
| coughing | Coughing voluntarily |
| deep breathing | deep inhale up to TLC, deep exhale to RV, no breathing for about 1-2 seconds, followed by deep inhalation to TLC and immediate exhalation to RV |
| humming | as “singing”, but with mouth closed (humming through the nose) |
| x s in, y s out | Breathing pattern with x seconds of deep inhalation and y seconds of deep exhalation in sync with the breathing visualisation video |
| 1/s exhalation | Breathing to a metronome set to 60 beats per minute, one exhalation per beat |
| deep breathing silent | as in “x s in, y s” out for x=10 s, y=8 s |
| deep breathing loud | as in “deep breathing silent” but vocalising “aah” throughout exhale |
| airways closure x s | deep inhale followed by x seconds of holding breath, then exhale to RV and repeat |
| wet-lips singing | as “singing” but wetting the lips by tongue in each pause |
| lip balm singing | as “singing” but with lip balm (Balea Lippenpflege Sensitive) put on the lips |
| lip balm shouting | as “shouting” but with lip balm (Balea Lippenpflege Sensitive) put on the lips |
| open-mouth shouting | as “shouting” but mouth kept open with lips not touching each other |
| open-mouth singing | as “singing” but mouth kept open with lips not touching each other |
| excessive \t\ articulation | speaking the consonant “t” at maximum loudness repeatedly with lips not touching each other |

Table S.2: Description of the respiratory activities investigated. The so-called *standard activities* done by all subjects were breathing, speaking normal, speaking loud and singing. The rest are referred to as *special activities* and are carried out to understand the origin and production mechanism of respiratory particles.

have an additional sealed barrier between the upper half of the face and the nose and mouth area. For both the new and the old isolation shield, the clean inhaled air enters the shield through a one-way inhalation valve on top of the shield. It then passes through two additional inhalation valves on each side of the nose into the sealed mouth/nose isolation shield area. When the subject exhales these valves are closed and the exhaled air enters the buffer volume (marked in light red in Fig. S.1) of the adaptor through one-way exhalation valves. In the new isolation shield (Fig. S.1**b**, the exhalation valve and buffer volume are directly on the bottom of the isolation shield. In the old isolation shield (Fig. S.1**c**), the air enters the mask's side volumes and flows up into the adaptor buffer volume. In both cases, the exhaled air is sampled with the PSS sampling tube from the adaptor and a filter is fixed on the exhale to prevent contamination of the surroundings/cleanroom with respiratory particles. The effectiveness of these setups is discussed in the following subsection S.1.4 and the influence of the adaptor buffer volume size is discussed in S.2.9.2

The sampled air from either **a** the new isolation shield, **b** the old isolation shield or **c** the funnel (see Fig. S.1), travels through PTFE tubes and passes through two diffusion dryers as described in the main paper. The samples are analysed in a combination of spectrometers: **d** TSI Optical Particle Sizer 3330 spectrometer (OPS) and TSI NanoScan Scanning Mobility Particle Sizer 3910 (SMPS) resulting in a sample flow rate of $1.75\,\mathrm{L\,min^{-1}}$ or **e** OPS, SMPS and TSI Aerodynamic Particle Sizer model 3321 (APS) resulting in a total flow rate of $6.75\,\mathrm{L\,min^{-1}}$. The details of the instrumentation are given in the main paper. When three PSS **e** were used the tubes were connected via the TSI flow splitter model 3708.

##### S.1.4 Validity of Sampling Methods

In the following, the sampling methods with funnel and isolation shield are discussed in more detail regarding potential non-isokinetic sampling and particle losses. At the funnel inlet with 15 cm diameter the sampling air speed is $0.17\,\mathrm{cm\,s^{-1}}$ or $0.64\,\mathrm{cm\,s^{-1}}$ depending on the combination of PSSs for OPS and SMPS (Fig. S.1**d**) or OPS, SMPS and APS (Fig. S.1**e**) respectively. With the funnel setup, no breathing experiments were performed, i.e. no nose breathing, therefore the exhalation flow can be assumed to be isoaxial. Losses due to impaction and non-isokinetic sampling because of flow direction can therefore be assumed to be minimal. To investigate potential non-isokinetic sampling with the funnel, we need to estimate the exhale air speed at funnel inlet. For this, we measured the exhaled air flow velocity for one subject at mouth height at a horizontal distance of 10 cm with a thermal anemometer probe (testo 405i). This distance is comparable to the distance between subject mouth and funnel in the setup shown in Fig. S.1**c**. For speaking normal, we found a mean velocity of about $10\,\mathrm{cm\,s^{-1}}$. This velocity is already in the range of typical turbulent fluctuations in a room. Moreover, we expect the exhale velocity to decrease with distance to the mouth in the cross-section. A different way to estimate exhale flow velocity at funnel distance would be to assume the exhale flow is uniform

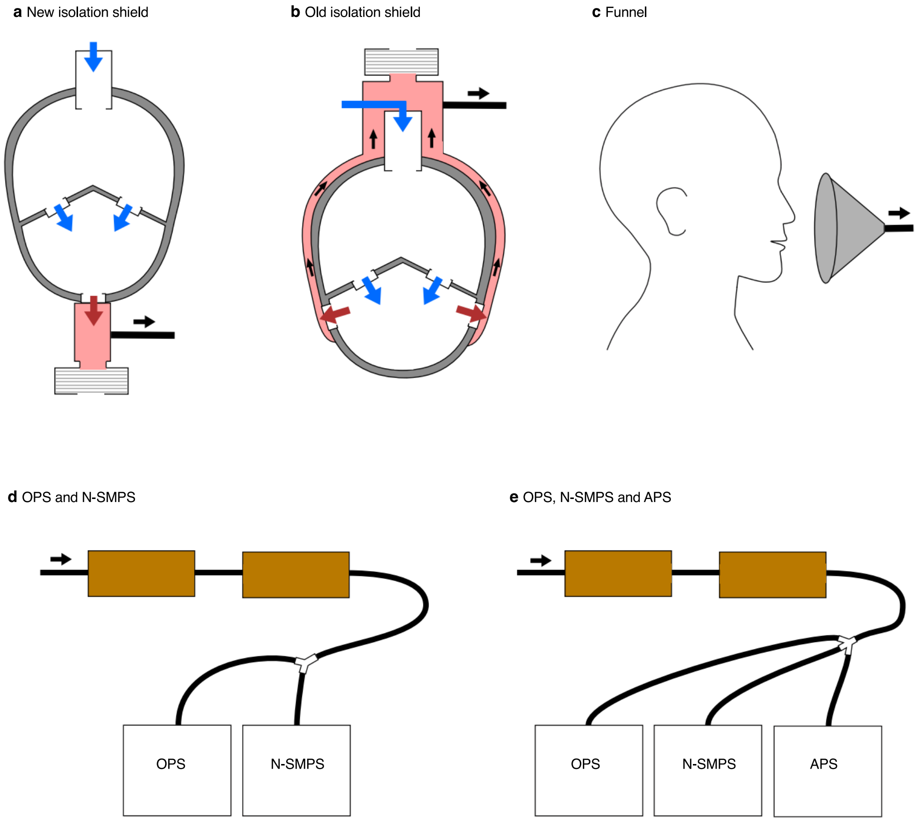


Figure S.1: First part of the measurement setup: sampling methods. To sample the exhaled air, either **a** the new isolation shield, **b** the old isolation shield or **c** a funnel was used. Blue arrows indicate valves that are only open during inhalation, red arrows indicate valves that are only open during exhalation. Shaded red areas represent the exhale buffer volume. The sampling tube is connected to one of the setups **d** or **e**. Second part of the measurement setup: the with one of the sample methods **a-c** sampled flow passes through PTFE tubes, two diffusion dryers and is then analysed in either **d** OPS and SMPS or **e** OPS, SMPS and APS.

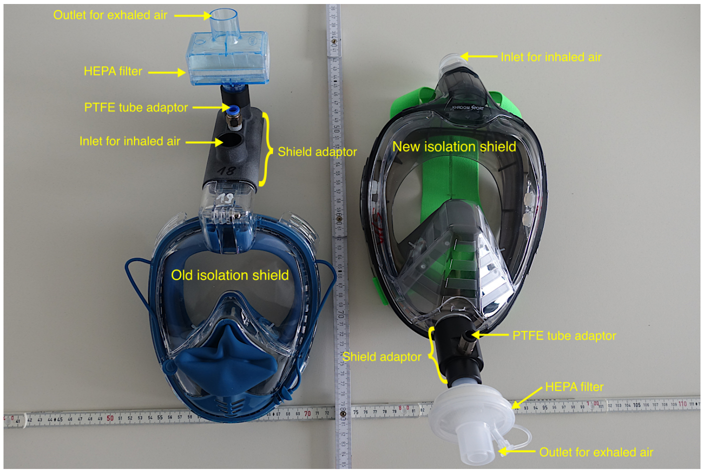


Figure S.2: Photograph of the old (left) and new (right) isolation shield used for sampling with the respective adaptors and exhalation filters attached.

**To obtain photos, please contact the corresponding author**

Figure S.3: Photograph of the different sampling strategies. **a** A subject is wearing the old isolation shield. A filter is both on the inhale inlet (optional in cleanroom) and exhale outlet (necessary) of the adaptor. **b** A subject is wearing the new isolation shield. A filter is put on the exhale outlet of the adaptor on the bottom of the shield. **c** Example of sampling with the holographic setup and the funnel. The sound pressure level, temperature and relative humidity are measured simultaneously.

and spans the whole cross-section of the funnel at the distance of 10 cm. The exhale flow velocity would be $1.26\,\mathrm{cm\,s^{-1}}$. Still assuming a uniform flow but parameters of [3] (mouth breathing) for mouth opening and exhale jet angles, the velocity at the funnel would be reduced to $0.32\,\mathrm{cm\,s^{-1}}$. Both calculated values are based on the volume flow rate of speaking loud measured on one subject ($13.4\,\mathrm{L\,min^{-1}}$, for singing $23.6\,\mathrm{L\,min^{-1}}$, not measured for speaking normal). With these estimations, we can assume the sampling with the funnel was performed sub-isokinetic which can lead to an overestimation of large particles or close to isokinetic. However, dilution of the exhaled air with the surrounding cleanroom air and therefore dilution of particle concentration is likely when sampling is performed with the funnel.

Comparing the Exhale Particle Size Distribution (EPSD) measured with the funnel to the EPSD measured with isolation shields (old and new, Fig. S.1**a**, **b**), shows that the isolation shield prevents this dilution. On average we sampled 2.6 times more PM5 with the isolation shield than we did with the funnel. In the shield, however, we have a stronger effect of impaction and sampling losses for >5 µm. This is caused by the exhalation valves and complex shield geometries (even more complex for the old mask) but also non-isokinetic sampling.

With the isolation shields the sampling is performed directly with the PTFE tube with 8 mm diameter inlet (tube itself has 6 mm inner diameter) from the exhale adaptor in a 90° angle to the exhalation flow. The sampling velocity is therefore $58.03\,\mathrm{cm\,s^{-1}}$ and $223.8\,\mathrm{cm\,s^{-1}}$ for the setups in Fig. S.1**d** and Fig. S.1**e** respectively. In the new shield, the flow of the exhalation goes through the cylindrical buffer volume (diameter 15 mm) with a speed of $122\,\mathrm{cm\,s^{-1}}$ (based exhalation volume flow rate measurement of breathing with $12.9\,\mathrm{L\,min^{-1}}$, similar for reading and even higher for singing). In the old shield, the flow through the buffer volume is slower as the cross-section is larger but also more complex and likely not laminar due to the complex shape of the flow path through the mask and the buffer volume adaptor. The high exhale flow velocity and the sampling at a 90° angle leads to undersampling of large particles i.e. particles with high inertia are less likely to follow the 90° redirected flow into the sampling tube. This also shows in the comparison between funnel sampled and isolation shield sampled data. Whereas for small particles the isolation shield performed better, for particles >5 µm the particle concentrations measured with funnel were larger and the difference further increased with particle size. We conclude that for small PM5 particles the isolation shield is the best sampling method since we have almost no dilution with surrounding air, for larger particles the funnel performs better due to fewer impaction losses and closer to isokinetic sampling. Therefore PM5 data measured with the funnel gets corrected by the ratio isolation shield/ funnel and concentrations of particles >5 µm are corrected with the funnel/ isolation shield ratio. For particles larger than ∼9 µm, all the PSS data collected by the funnel or the isolation shields is replaced with holographic data which also prevents any influence of the potential sub-isokinetic sampling that would lead to an overestimation of large particles in the funnel data and losses within the sampling tubes, which are most significant for large particles. Moreover, the holographic samples are less likely to be diluted as they were taken at <5 cm in front of the subjects' mouths. In all cases, independent of sampling strategy (with PSSs) the long tubes with partially high curvatures and the two dryers are associated with particle losses. A correction is applied for this as described in the methods section of the main paper. Since these losses are independent of sampling methods it does not affect the discussion above. For the two dryers, we found an average loss rate of 19 % in an inter-comparison experiment using dolomite dust at concentrations up to 350 $cm^{-3}$. The loss rate was nearly independent of particle size and concentration in a concentration range from 8 $cm^{-3}$ $\leq N \leq$ 350 $cm^{-3}$.

##### S.1.5 Shrinkage Factor

The shrinkage factor of exhaled particles is investigated experimentally. In the first approach exhale particle concentration is measured in the cleanroom with and without the diffusion dryers on 11 subjects (200 experiments totalling 370 minutes of measurements) during breathing and singing. For "direct" measurements without the diffusion dryers, the subjects exhaled or sang directly and at a distance of a few centimetres from the sampling inlets of the OPS or the APS. Fig. 2 shows the ratio of particle size measured without the dryers to the particle size with the same concentration when the dryers are used. It can be seen that most sub-micron particles are fully dried, i.e. detected shrinkage $\sim 1$, even when they are exhaled directly to the sampling inlets. For particles >3 µm a plateau is reached which suggests that these particles could shrink in size by a factor of 4.5 when passed through the diffusion dryers. This figure also shows the risk of measuring exhale particles without controlling the RH, which leads to mixing concentration of fully dried, partly dried and non-dried particles with each other.

In the second approach, we measured the shrinkage factor and the rate of evaporation of 9 saliva droplets from one subject and in total 36 ASL (airway surface liquid) samples from four subjects. For this, the droplets were suspended each on a single horizontal strand of human hair with a mean diameter of 50 µm and let to dry. The relative humidity of the room was $\leq$ 30% for most experiments (if not explicitly stated otherwise) and the temperature was 23.1 °C to 23.2 °C. This method of suspension ensures that the whole droplet surface is exposed to the surrounding air and the humidity gradient is similar to that around a freely floating droplet, unlike in the case of a drop resting on a surface. In order to prevent air drafts from affecting the evaporation rate, we enclosed the experiment in a large transparent glass box. We recorded the silhouette of the drying droplets using a Phantom VEO4K 990L camera with the optical axis oriented horizontally and perpendicular to the hair. We used an approximate method to measure the droplet size at any time, which rests on the approximation that the cross-section of the droplet in the plane normal to the hair is roughly circular in shape. One can then obtain the droplet volume by simply integrating the squared apparent diameter along the hair length and subtracting the contribution due to the hair itself. The approximation is certainly valid in the initial stages of the drying process, when the droplet equivalent spherical diameter is much larger than the hair diameter, since the droplet was close to spherical (Bond number was at most 0.2). In the later stages, this approximation might break down due to increased effects of impurities and small volume. In order to check the cross-section shape of the dry residual, we rotated the hair to nearly align with the camera optical axis and confirmed that it was indeed typically a nearly circular ellipse. As a further check of the volume-computing method, we selected three dry residuals at random and obtained their convex hulls by rotating the hair with the residual along its axis and recording the silhouette as before. The equivalent spherical diameters of the residuals computed from the convex hulls were 3, -5 and 14% different from those obtained by the circular cross-section approximation. Figure S.4 shows the obtained equivalent spherical diameter D(t) as a function of time normalised with the equilibrium dry diameter for both saliva and ASL droplets. Saliva droplets were sampled either after at least 15 min without drinking (no water) or drinking a sip of water immediately before the experiment (drinking water before). During some of the saliva droplet measurements, the ambient relative humidity was relatively high (45%), which affected the evaporation rate. The verified ASL droplets are marked by opaque, dotted lines.

##### S.1.6 The effect of diffusion dryers on relative humidity within the experimental setup

The relative humidity inside the particle sampling system was measured with one subject for three standard activities, which were breathing normally, reading at normal voice, and singing "Happy Birthday". Between the diffusion dryers and the PSS, a Vaisala HMP7 temperature and humidity sensor was placed to measure temperature $T$ and relative humidity RH in a distance from the isolation shield where usually the PSSs would be located. The first experiment was done without diffusion dryer, the second experiment was done with one diffusion dryer (type: see description of the setup) and the third experiment was done with both diffusion dryers as usually. Experiment 1 acts as a reference as it shows what would happen inside the tubing if the sample flow was not dried. Ambient conditions in the cleanroom were $T = 21\,°C$ and RH of 44 %. The first observation is that it took 115 s from the beginning of the breathing experiment until RH changed. Within less than 3 min, RH exceeded 85 %. The maximum RH of 87.5 % was reached 220 s after the end of the breathing experiment. 8 min after the end of the breathing experiment, RH was decreasing rapidly, following an exponential decay. The overall trend did not change when comparing the data from breathing with reading or singing. Fig. S.5 shows the time series data of RH for the three breathing experiments with 0, 1 and 2 dryers.

With one diffusion dryer, the curves still look similar, but the maximum RH is much lower than in the case without diffusion dryer. Here, the ambient RH was as low as 12 % and even after 5 min of breathing, the maximum (which arrived 315 s after the end of the experiment) was at RH of 19 %. This experiment has shown that even one diffusion dryer reduces RH inside the particle sampling system to values <20 %, which means that the water must have

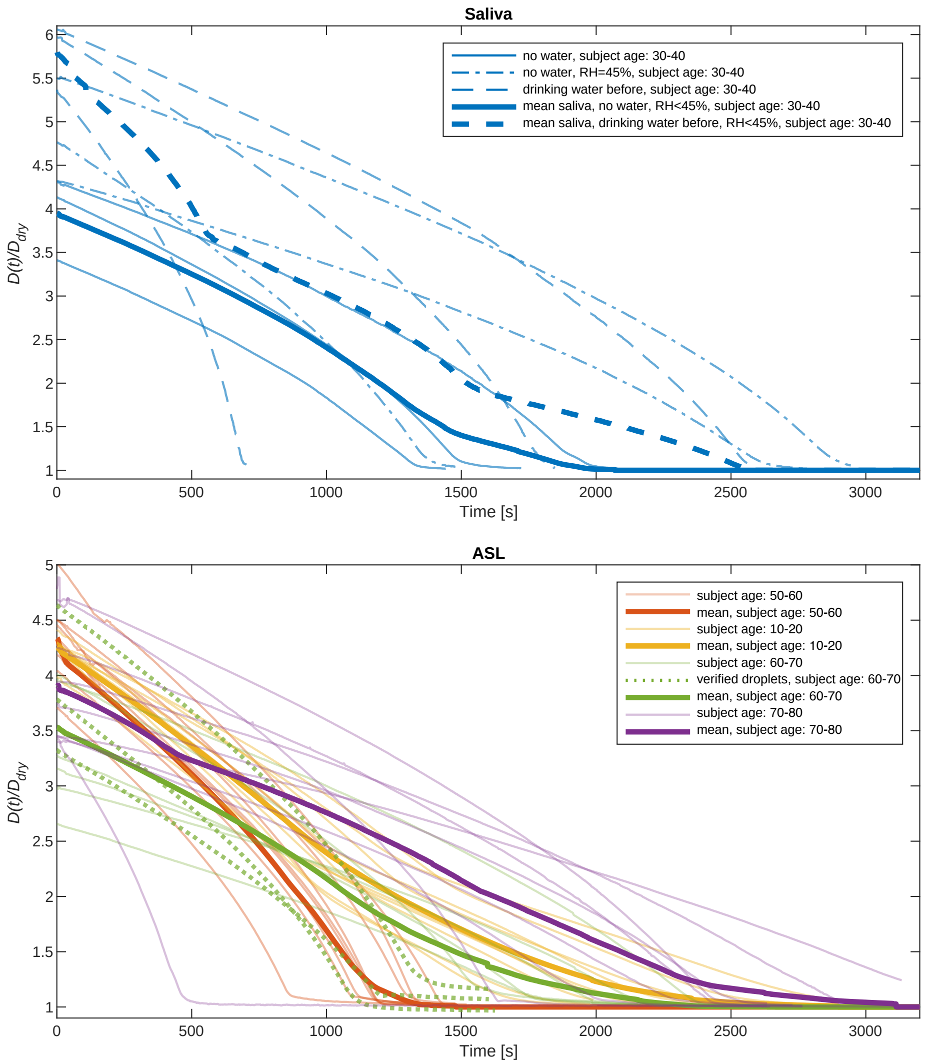


Figure S.4: Equivalent spherical diameter of the 9 analysed saliva and 36 ASL particles as a function of time. The results are shown for each particle individually and the mean of each subject is shown. The diameter is normalised with the equilibrium dry diameter. Verified particles are the particles for which the volume calculation via circular cross-section approximation was validated by measuring the convex hulls of the dry residual.

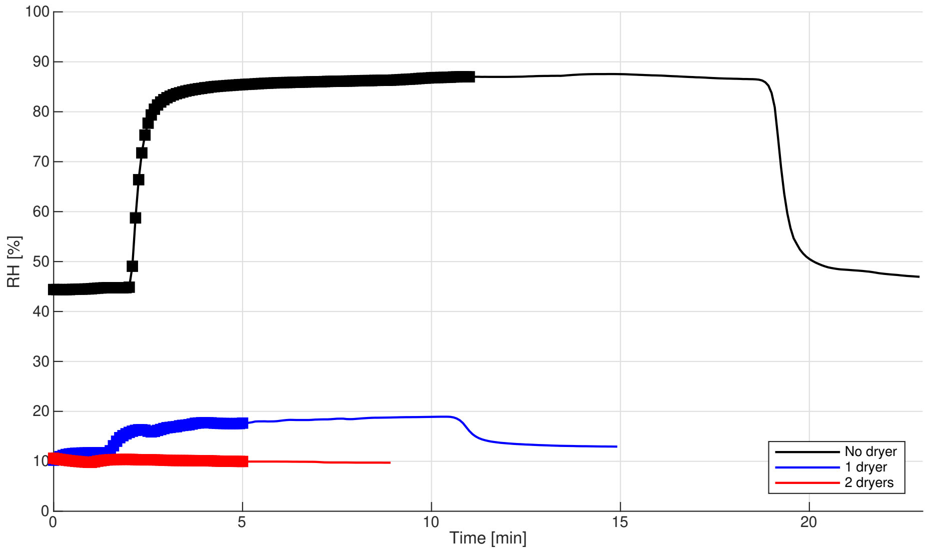


Figure S.5: RH time series of three breathing experiments with no diffusion dryer (black), one diffusion dryer (blue) and two diffusion dryers (red). The squares represent the time when the subject was doing the breathing experiment.

been fully evaporated from the exhaled particles before they reached the particle spectrometers. The only thing that can explain the late decrease of RH after the end of the breathing experiments is condensed water in the tube between the isolation shield and the first diffusion dryer.

At ambient conditions of $T = 21\,^{\circ}\mathrm{C}$ and RH$= 44\,\%$, it took 315 s from the end of the second experiment to evaporate the water inside the tube. During that time, an air volume of approximately 9.2 L went through the tubing. When considering the actual and saturation water vapour partial pressure and the volume of air needed to start the decrease in measured RH, we estimate the amount of liquid water in the tubing to be on the order of 0.1 g.

In the experiment with the two dryers, the time between setup and start of the experiment was too short, so that the ambient RH was not reached. Right before the start of the breathing experiment, a local minimum of RH was found at 9.8 %. However, RH decreased even more throughout the experiment, with a final value below 9.4 %.

Even with the usage of an ultrasonic nebuliser, it was not possible to see an increase in RH during the experiment when using two diffusion dryers.

##### S.1.7 Calibration of particle size in the holographic setup

The holographic setup was calibrated with NIST traceable calibration glass microspheres from ThermoFisher Inc. in seven different sizes from 8 to 50 µm diameter. The particles were released manually into the sample volume where they were visible for about a second as a thin cloud. Using a particle dispenser with a propellant is not recommended for holography as the difference in density and/or temperature between the propellant and the ambient air will create schlieren which make the automated reconstruction and particle detection impossible [4]. Here, we applied the same restriction of the sample volume as for the exhaled droplets. The exact same method as for the respiratory particles was used to estimate the particle diameter. Overall, there were only two bead sizes out of the tolerance by 1.0 µm at maximum, the rest was well within the tolerance. Fig. S.6 shows the cumulative distribution functions of the measured and nominal particle diameters. The fact that the nominal and measured standard deviations do not deviate that much from each other shows that the nominal size distributions are pretty much in agreement with the measurements. The two outliers are the smallest beads (7.7 µm nominal diameter) and the 20 µm beads.

| $\mu_n$ [µm] | $\sigma_n$ [µm] | $\mu_H$ [µm] | $\sigma_H$ [µm] | dev. [µm] | dev. tol. [µm] |
|---|---|---|---|---|---|
| 7.7 ± 0.4 | 1.0 | 9.1 | 1.4 | +1.4 | +1.0 |
| 9.8 ± 1.0 | 1.2 | 10.0 | 1.2 | +0.2 | within |
| 14.4 ± 0.8 | 1.8 | 13.9 | 1.5 | -0.5 | within |
| 20.2 ± 1.2 | 2.8 | 18.3 | 2.4 | -1.9 | -0.7 |
| 29.5 ± 1.0 | 1.9 | 28.9 | 2.3 | -0.6 | within |
| 42.3 ± 1.1 | 1.5 | 42.3 | 1.6 | 0 | within |
| 49.3 ± 1.4 | 3.4 | 48.7 | 2.2 | 0.6 | within |

Table S.3: Nominal mean particle diameter $\mu_n$ with errorbars given by the manufacturer, nominal standard deviation $\sigma_n$ and mean particle diameter $\mu_H$ and standard deviation $\sigma_H$ measured by the holographic setup for the calibration with glass microspheres. Also the deviation between nominal and measured mean diameter, and the deviation from the tolerance interval (if applicable) is listed.

##### S.1.8 Parameters of Multimodal log-normal fit

Table S.4 shows fit parameters found for the multimodal log-normal fit to the subjects-averaged particle size distribution. For all activities except shouting, the mode diameters and geometric standard deviation are set in an iterative process aimed to minimise the number of modes required to represent the data for all activities, while the amplitudes are found by minimising the difference between the fit and the data points for each activity. For shouting the modes, geometric standard deviation and amplitudes are allowed to change during the fitting procedure. The elevated concentration seen at 4.6 µm for all the activities is an instrument bias caused by ripples in Mie scattering profiles of the PSS and

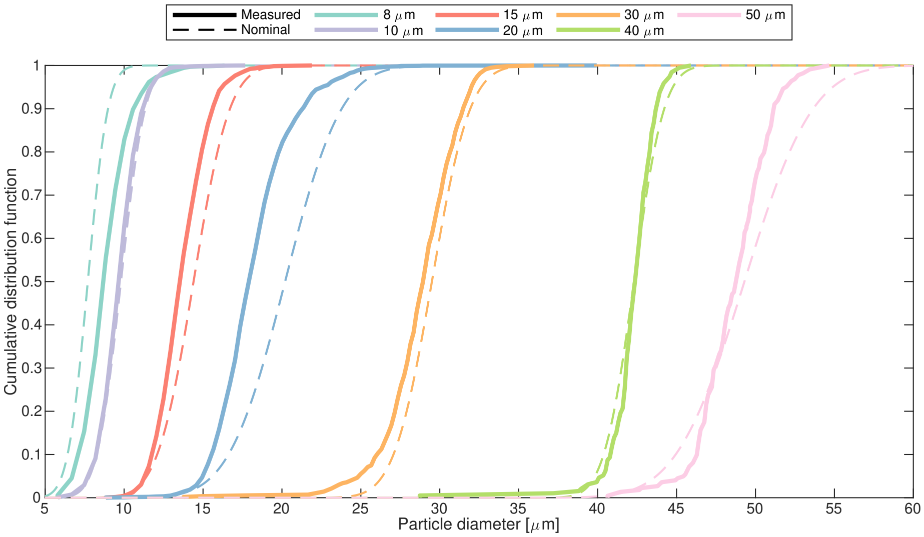


Figure S.6: Cumulative distribution functions of particle diameter measured for glass spheres used as calibration standard (thin dashed lines), and actual particle sizes measured with the holographic setup (thick solid lines). The sample size for each calibration run was 194 particles or more.

the choice of bin cut-points, and is not related to the physics of exhale emissions. Therefore the log-normal fits are intentionally not fitted to this channel but to the three-point geometric mean.

#### S.2 Supplementary Results

In the following, a more detailed look into parameters potentially affecting EPSD is presented. For this, only the results measured with the OPS are taken into account (0.3-10 µm dry diameter, corresponding to exhaled diameter $D_0$ =1.36-45.5 µm). When not indicated otherwise, the particle diameter is given as the exhaled diameter, $D_0$. When size dependency is not shown the quantities total emitted particle number concentration of PM5, $N_{<5}$, and total emitted particle volume concentration of PM5, $V_{<5}$, are shown. $N_{<5}$ is the total counted particles in all OPS bins up to 5 µm exhaled diameter per sample volume and $V_{<5}$ the total volume of these particles (assuming spherical particles) before drying per sample volume. In the following, low-emitters are defined as those whose exhalation concentration is one geometric standard deviation below the geometric mean concentration, and super-emitters are defined as those whose exhalation concentration is one geometric standard deviation above the geometric mean concentration of the population.

| activity | $i$ | | | | | | | $R^2$ |
|---|---|---|---|---|---|---|---|---|
| | $1^*$ | 2 | 3 | 4 | 5 | $6^*$ | $7^*$ | |
| $d_i$ [$\mu m$] | 0.05 | 0.15 | 0.56 | 9.0 | 38 | 195 | 700 | |
| $\sigma_i$ | 0.38 | 0.60 | 1.0 | 0.85 | 0.70 | 0.70 | 0.60 | |
| | $A_1$ [cm$^{-3}$] | $A_2$ [cm$^{-3}$] | $A_3$ [cm$^{-3}$] | $A_4$ [cm$^{-3}$] | $A_5$ [cm$^{-3}$] | $A_6$ [cm$^{-3}$] | $A_7$ [cm$^{-3}$] | |
| breath. | 0.37 | 0.74 | 1.08 | 0.006 | - | - | - | 0.96 |
| speak. norm. | 1.26 | 2.19 | 3.39 | 0.04 | 0.22 | 0.01 | - | 0.99 |
| speak. loud | 3.0 | 4.5 | 6.4 | 0.09 | 0.22 | 0.006 | 0.003 | 0.99 |
| singing | 2.96 | 7.18 | 8.27 | 0.14 | 0.21 | 0.004 | - | 0.99 |
| $d_i$ [$\mu m$] | 0.05 | 0.12 | 0.49 | 8.8 | 31 | 146 | - | |
| $\sigma_i$ | 0.29 | 0.50 | 1.1 | 0.65 | 0.58 | 0.35 | - | |
| | $A_1$ [cm$^{-3}$] | $A_2$ [cm$^{-3}$] | $A_3$ [cm$^{-3}$] | $A_4$ [cm$^{-3}$] | $A_5$ [cm$^{-3}$] | $A_6$ [cm$^{-3}$] | $A_7$ [cm$^{-3}$] | |
| shouting | 0.9 | 43.3 | 43.3 | 0.63 | 0.46 | 0.009 | - | 0.95 |

Table S.4: Fit parameters found for different respiratory activities carried out in this study. For each activity the bin-normalised number concentrations is fitted with a multimodal lognormal equation $\mathrm{d}N/\mathrm{dLog}D_0 = \sum_{i=1}^{n} A_i \cdot \exp\left(-\left(\ln\left(D_0/d_i\right)/\sigma_i\right)^2\right)$, where $A_i\,[cm^{-3}]$ is the mode $i$ amplitude, $d_i\,[\mu m]$ is the mode $i$ diameter, $\sigma_i$ is the mode $i$ geometric standard deviation and $D_0\,[\mu m]$ is the particle diameter at the exhalation. It should be noted that the $1^*$, $6^*$ and $7^*$ modes are fitted to a few data points close to the detection limit of the instruments. Furthermore, data points close to $6^*$ and $7^*$ modes are associated with large uncertainties. Therefore, the presented fitting parameters are only valid for the size range from 50 nm to 1000 µm while considering large uncertainties for >100 µm particles.

##### S.2.1 Effect of voice sound pressure

The A-weighted-decibels dBA 3rd quartiles measured at a distance of ~20 cm away from the subject for vocalisation activities averaged over all the subjects are 78.6 dBA, 83.6 dBA, 85.7 dBA and 102.4 dBA for speaking normally, speaking loudly, singing and shouting respectively. This suggests that the elevated particle concentration, as an example, from speaking loudly to speaking normally is most likely due to an increase in sound pressure. There is a connection between loudness and being a low-emitter or a super-emitter. For speaking normal, we found a mean of the loudness 3rd quartile of 77.6 dB(A) in the cohort of the low-emitters, and 82.7 dB(A) in the cohort of the super-emitters. The difference between the two cohorts was not significant (p = 0.083). For speaking loud, we found a mean of the loudness 3rd quartile of 81.1 dB(A) in the cohort of the low-emitters, and 89.5 dB(A) in the cohort of the super-emitters, which is a significant difference between the two cohorts (p = 0.006). The biggest difference was found for singing with a mean of 3rd quartile loudness of 81.2 dB(A) in the low-emitters and 92.0 dB(A) in the super-emitters, which was also significant ($p = 6 \cdot 10^{-5}$). In the case of speaking normal, the standard deviation of 3rd quartile loudness was 5.1 dB(A) in the super-emitters and 7.9 dB(A) in the low-emitters. For speaking loud, the standard deviation of 3rd quartile loudness was 6.2 dB(A) in the super-emitters and 5.2 dB(A) in the low-emitters. For singing, the standard deviation of the 3rd quartile loudness was 5.6 dB(A) in the super-emitters and 5.1 dB(A) in the low-emitters. This means that the difference of the mean 3rd quartile loudness for singing in both groups is about two standard deviations large. So the difference between low-emitters and super-emitters in the phonetic activities can at least partially be explained by the loudness.

However, each subject had a different increase in the sound pressure from speaking normally to loudly – among other variable parameters. To investigate the impact of sound pressure itself, we have constrained our analyses to 76 subjects aged between 5-75 years who have performed speaking normally, speaking loudly and singing. For these subjects we found that there is a significant correlation, i.e. $p < 0.01$, between the concentration of 1.5-7 µm and sound pressure as shown in Fig. S.7. Maximum correlation of $R^2 = 0.51$ between sound pressure and particle concentration is found for ~2.5 µm particles. There is, however, a significant scatter in the data, which lead to low $R^2$ values between the linear regression and the measurements. Furthermore, the change in the particle concentration is well within the within-subject variability for a fixed activity.

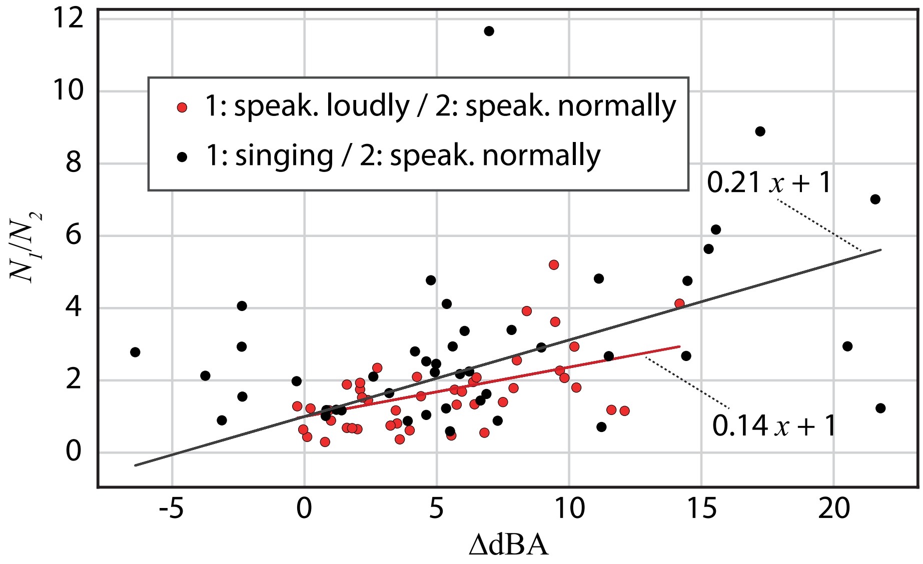


Figure S.7: Impact of change in sound pressure in the EPSD concentration for the same subject when loudly reading a phonetic-standard text or singing “happy birthday” to when they are reading the phonetic-standard standard text with normal voice for particles with 1.5 µm< $D_0$ <7 µm.

##### S.2.2 Subject Variability

The within-subject variability is shown in Fig. S.8 for the five subjects with the highest number of total independent measurements. It spans up to one full order of magnitude for both PM5 particle number concentration and volume concentration. When comparing the within-subject variability for different activities and subjects (S.8) the different numbers of independent-tests $n$ have to be taken into account. The independent measurements of these subjects were performed over the span of 200 days, but some also on the same day. The exhaled particle concentration of each subject can vary as much in a single day as it does over a longer time span up to over 200 days. For Subject 20, the number of experiments for breathing and singing was large enough to determine the distribution type. In case of $N_{<5}$ for breathing (18 experiments), we could fit a Gaussian distribution with $\mu = 0.206\,\mathrm{cm}^{-3}$, $\sigma = 0.164\,\mathrm{cm}^{-3}$ and $R^2 = 0.937$, and for singing (14 experiments), we could fit a Gaussian with $\mu = 0.392\,\mathrm{cm}^{-3}$, $\sigma = 0.205\,\mathrm{cm}^{-3}$ and $R^2 = 0.967$. The other activities and subjects did not contain enough experiments for a reasonable fit. Both Gaussian distributions we could fit to the data are relatively broad, which means that the standard deviation is 50 % of the arithmetic mean value or more than that ($\sigma/\mu = 0.796$ and $\sigma/\mu = 0.523$). For $V_{<5}$, we also get good fits to a Gaussian distribution (Breathing: $\mu = 1.60\,\mathrm{\mu m^3\,cm^{-3}}$, $\sigma = 1.19\,\mathrm{\mu m^3\,cm^{-3}}$ and $R^2 = 0.945$, Singing: $\mu = 2.96\,\mathrm{\mu m^3\,cm^{-3}}$, $\sigma = 1.51\,\mathrm{\mu m^3\,cm^{-3}}$ and $R^2 = 0.956$). The $\sigma/\mu$ ratios are $\sigma/\mu = 0.698$ and $\sigma/\mu = 0.510$ respectively.

The between-subject variability for the standard activities is shown in Fig. S.9. The variability in total PM5 number and volume concentration spans up to 2.5 orders of magnitude. When comparing the between-subject variability for different activities (Fig. S.9) the different number of subjects that performed each activity has to be taken into account. Table S.5 shows the arithmetic and S.6 the geometric mean and standard deviation of $N_{<5}$ and $V_{<5}$ as another measure of between-subject variability. As shown in Subsection S.2.8, we found a log-normal distribution for $N_{<5}$ and $V_{<5}$, which is the reason for presenting the *geometric* mean and standard deviation in Table S.6. While cumulative quantities like $N_{<5}$ and $V_{<5}$ were always non-zero for the entire cohort of subjects and all experiments, this condition is not necessarily fulfilled for $N$ in individual size channels of the particle size spectrometers. Because of that, we could only investigate the distribution type for $N_{<5}$ and $V_{<5}$ and not for each size bin individually and when particle size distributions are investigated the arithmetic mean was used (e.g. Figure 1, main paper). The arithmetic mean and standard deviation as well the ratio of standard deviation and mean are given in Table S.5 for completeness and to also give a more intuitive measure of the scatter of the data. How the between-subject variability changes, when only subjects of a certain age subgroup are considered, is shown in Table S.7 where the values of $\mu_g$ and $\sigma_g$ were obtained from log-normal fits instead of direct calculation from the data with the intention to get more robust estimates of $\mu_g$ and $\sigma_g$ even if the number of subjects is small. Figure S.10 and S.11 show the between-subject variability for the number concentrations measured in the particle size ranges of the OPS bins individually.

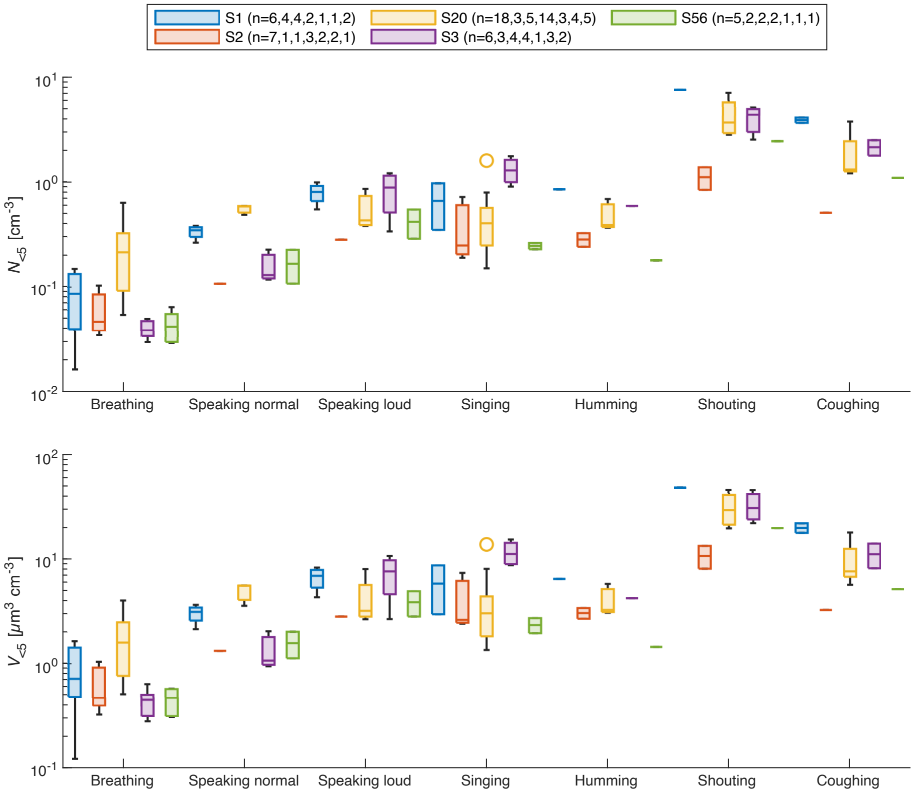


Figure S.8: Within-subject variability for the five subjects with the highest number of independent measurements in the standard activities for both $N_{<5}$ and $V_{<5}$. The number of independent measurements $n$ can be found in the legend for each subject in the order: breathing, speaking normal, speaking loud, singing, humming, shouting, coughing. Only measurements where sampling was performed with either the new or old isolation shield are considered.

##### S.2.3 Respiratory particle emission vs. gender

When comparing the total exhaled PM5 particle number concentrations in the activities presented in the main paper (breathing, speaking normal, speaking loud, singing, humming shouting, coughing) for male and female subjects we see slightly higher particle concentrations for male subjects. However, it has to be taken into account that gender of subjects was unevenly distributed in the age groups. To disentangle the effect of age and gender Figure S.12 shows a scatter plot of the mean PM5 particle exhalation per activity and subject. The results of female subjects are presented in red, the results of male subjects in

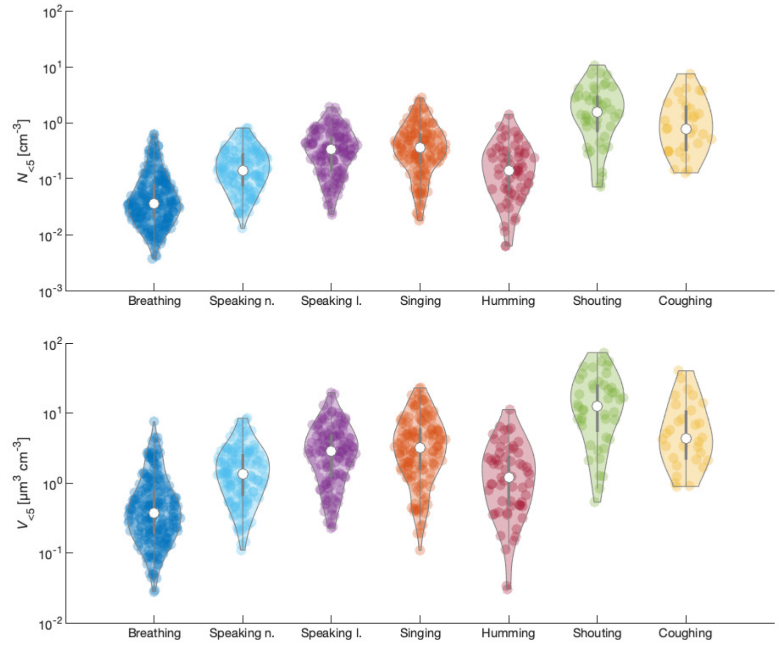


Figure S.9: Between-subject variability for the standard activities. The scatter in exhaled number concentration $N_{<5}$ of PM5 particles and in the total exhaled volume concentration $V_{<5}$ of PM5 particles is shown via a violin plot. If subjects performed more than one experiment per activity the mean is shown. Only measurements were sampling was performed with either the new or old isolation shield are considered. The plot was made with [2].

| activity | $\mu\,(N_{<5})$ [cm$^{-3}$] | $\sigma\,(N_{<5})$ [cm$^{-3}$] | $\frac{\sigma(N_{<5})}{\mu(N_{<5})}$ | $\mu\,(V_{<5})$ [µm$^3$ cm$^{-3}$] | $\sigma\,(V_{<5})$ [µm$^3$ cm$^{-3}$] | $\frac{\sigma(V_{<5})}{\mu(V_{<5})}$ |
|---|---|---|---|---|---|---|
| Breathing | 0.06 | 0.08 | 1.34 | 0.66 | 0.96 | 1.45 |
| Speaking n. | 0.19 | 0.17 | 0.88 | 1.76 | 1.64 | 0.93 |
| Speaking l. | 0.26 | 0.35 | 1.34 | 3.60 | 3.44 | 0.96 |
| Singing | 0.49 | 0.50 | 1.02 | 4.65 | 4.72 | 1.02 |
| Humming | 0.22 | 0.25 | 1.15 | 1.79 | 2.02 | 1.13 |
| Shouting | 2.16 | 2.36 | 1.09 | 17.11 | 17.40 | 1.02 |
| Coughing | 1.27 | 1.68 | 1.32 | 7.35 | 9.91 | 1.35 |

Table S.5: Arithmetic mean $\mu$ and standard deviation $\sigma$ and their ratio for either the PM5 particle number concentration $N_{<5}$ or the PM5 exhaled particle volume concentration $V_{<5}$.

| activity | $\mu_g\,(N_{<5})$ [cm$^{-3}$] | $\sigma_g\,(N_{<5})$ | $\mu_g\,(V_{<5})$ [µm$^3$ cm$^{-3}$] | $\sigma_g\,(V_{<5})$ |
|---|---|---|---|---|
| Breathing | 0.04 | 2.64 | 0.37 | 2.78 |
| Speaking normal | 0.13 | 2.45 | 1.19 | 2.53 |
| Speaking loud | 0.26 | 2.63 | 2.33 | 2.72 |
| Singing | 0.29 | 3.05 | 2.68 | 3.21 |
| Humming | 0.12 | 3.33 | 0.98 | 3.43 |
| Shouting | 1.16 | 3.56 | 9.60 | 3.40 |
| Coughing | 0.69 | 3.03 | 3.99 | 2.91 |

Table S.6: Geometric mean $\mu_g$ and geometric standard deviation $\sigma_g$ for either the PM5 particle number concentration $N_{<5}$ or the PM5 exhaled particle volume concentration $V_{<5}$.

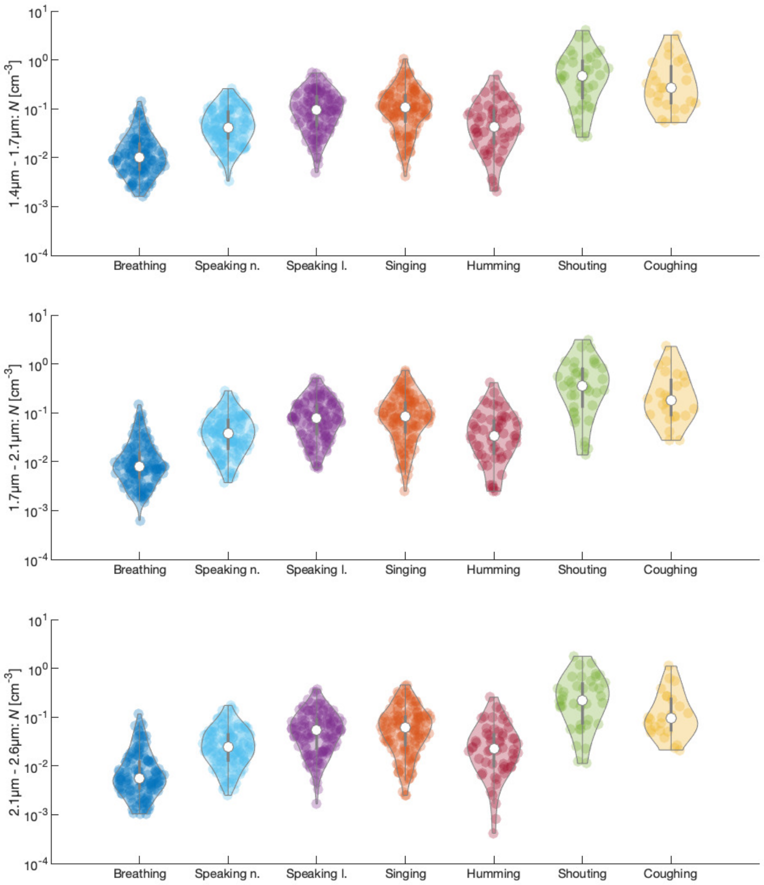


Figure S.10: Variability in total exhaled particle concentration between different subjects for each OPS bin individually. The particle diameter range given in the y-axis label is expressed in exhaled diameter $D_0$. The plot was made with [2].

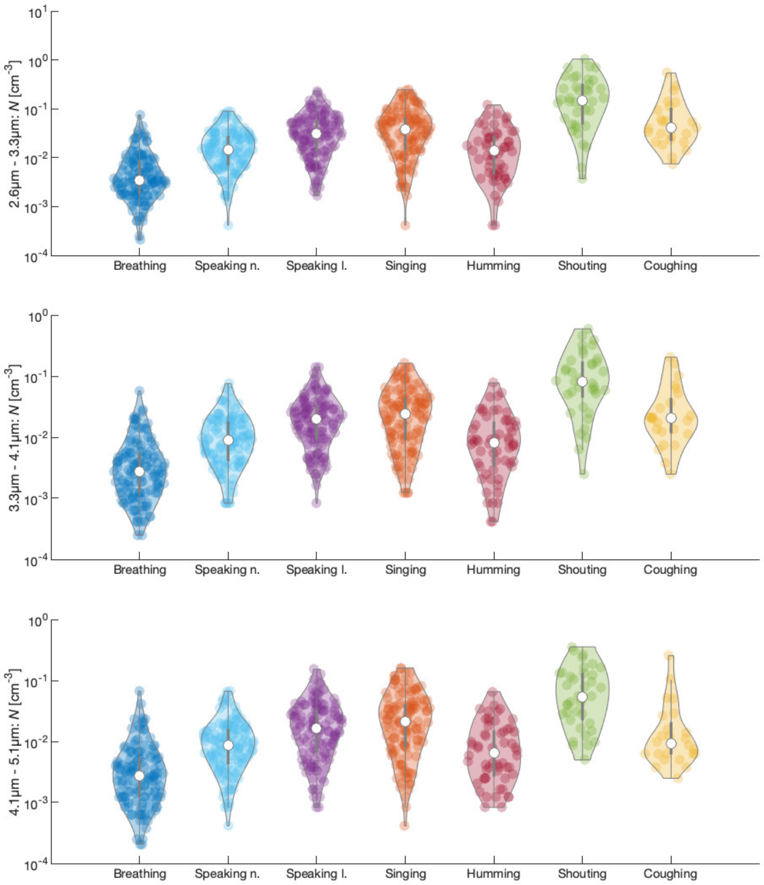


Figure S.11: Variability in total exhaled particle concentration between different subjects for each OPS bin individually. The particle diameter range given in the y-axis label is expressed in exhaled diameter $D_0$. The plot was made with [2].

blue. We can see that both genders follow the same trend with age and more importantly, that the PM5 exhaled particles of female subjects cover the same range as the ones of male subjects in the respective age. We observe the same in individual scatter plots for particles in each size bin individually and analysis of comparing female and male EPSD for different age groups also did not show any trends between the genders in any of the activities. We can therefore conclude that gender plays no significant role in particle production/exhalation.

##### S.2.4 Respiratory particle emission vs. smoking

As each subject was also asked about being a smoker or a non-smoker, we examined possible differences in the respiratory particle emission similar to the physical activity. Here, we have the issue that only 7 subjects were smokers. To make the comparison more representative, we included only those subjects for comparison who were in the same 5-year age group as the smokers (20-24 years, 25-29 years, 45-49 years, 55-59 years, 60-64 years), which gives us 28 subjects total for the control group. Qualitatively, we see that three out of the seven smokers had the highest emitted number concentration for their age group in breathing. For speaking normal and speaking loud, one of seven smokers was the highest in their age group. For singing, all smokers were below one or more non-smokers in terms of emitted number concentration. When performing a Student's t-test on $\log(N_{<5})$ and $\log(V_{PM5})$, we could not find a statistically significant difference between smokers and non-smokers. The result of the t-test is the same for $N_{\mathrm{PM10}}$ and $V_{\mathrm{PM10}}$.

##### S.2.5 Respiratory particle emission vs. exercise habits

In our questionnaire, we asked each subject how many whole hours per week they exercise of whatever kind. Figure S.14 shows a box plot of $N_{<5}$ and $V_{<5}$ for the activities breathing, speaking normal, speaking loud and singing for groups with different exercise habits. Among all subjects (including children and teens) we found a median of 3 hours per week. No clear trend is observable. To further investigate a possible statistical significance of exercise habits on the respiratory particle emission, we subdivided the whole sample in an "inactive" group (exercise hours per week less or equal to the median) and an "active" group (at least 4 hours of sports per week). On average we had 64 subjects in the inactive group per activity, and 54 subjects in the active group. If a subject did the same activity multiple times on different days, we took the median of $N_{<5}$ and $V_{<5}$ from this subject. As we show in Subsection S.2.8 that $N_{<5}$ and $V_{<5}$ follow a log-normal distribution, we performed a double-sided Student's t-test for $\log(N_{<5})$ and $\log(V_{PM5})$ for each of the standard activities breathing, speaking normal, speaking loud, and singing. In one of the cases ($N_{<5}$ for speaking loud) we found a statistically significant difference between the active and the inactive groups ($p = 0.03$) but not for any other activity or for $V_{<5}$. As the correlation between aerosol emission and age is strongest for PM5, this observation might actually originate from age rather than exercise habits. The

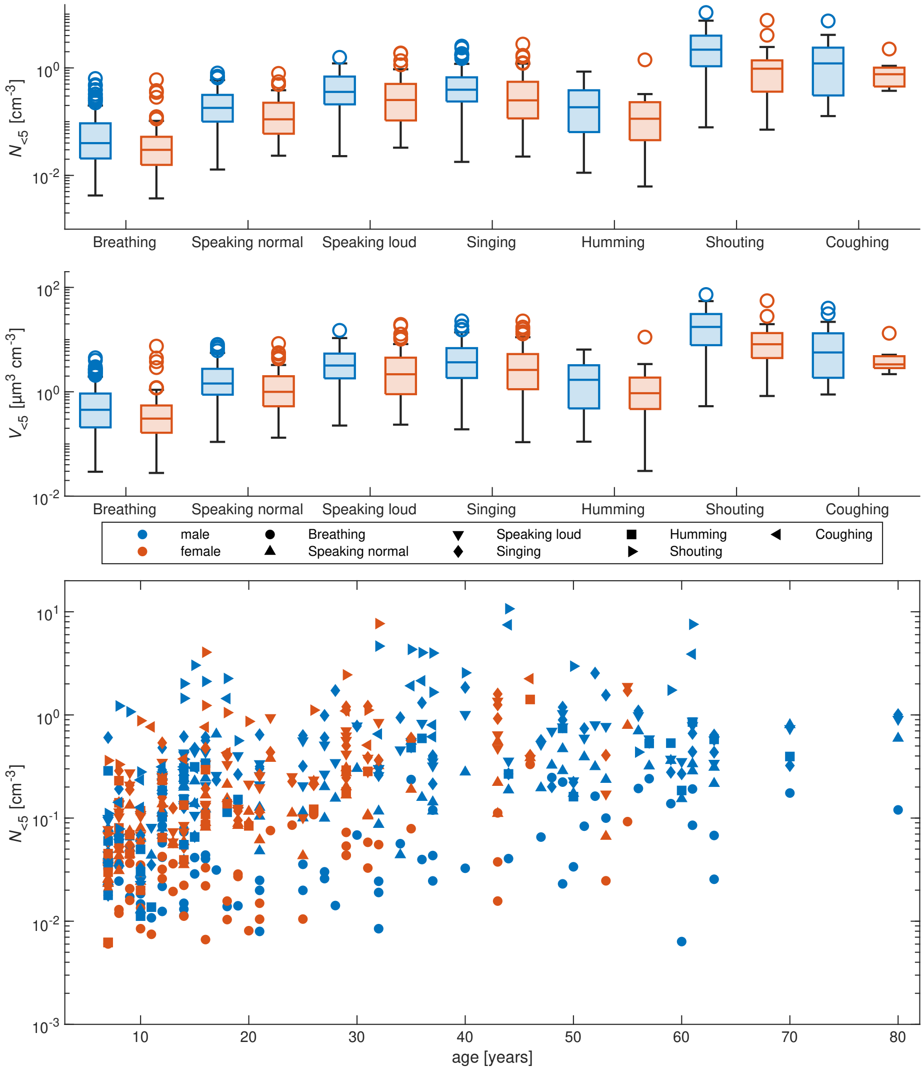


Figure S.12: Top: Total exhaled PM5 particle concentrations compared for male and female subjects. The comparison is shown for the standard activities established in the main paper. Bottom: Scatter plot of total exhaled PM5 particle concentration $N_{<5}$ per subject and activity as a function of age marked for male subjects in blue and in red for female subjects.

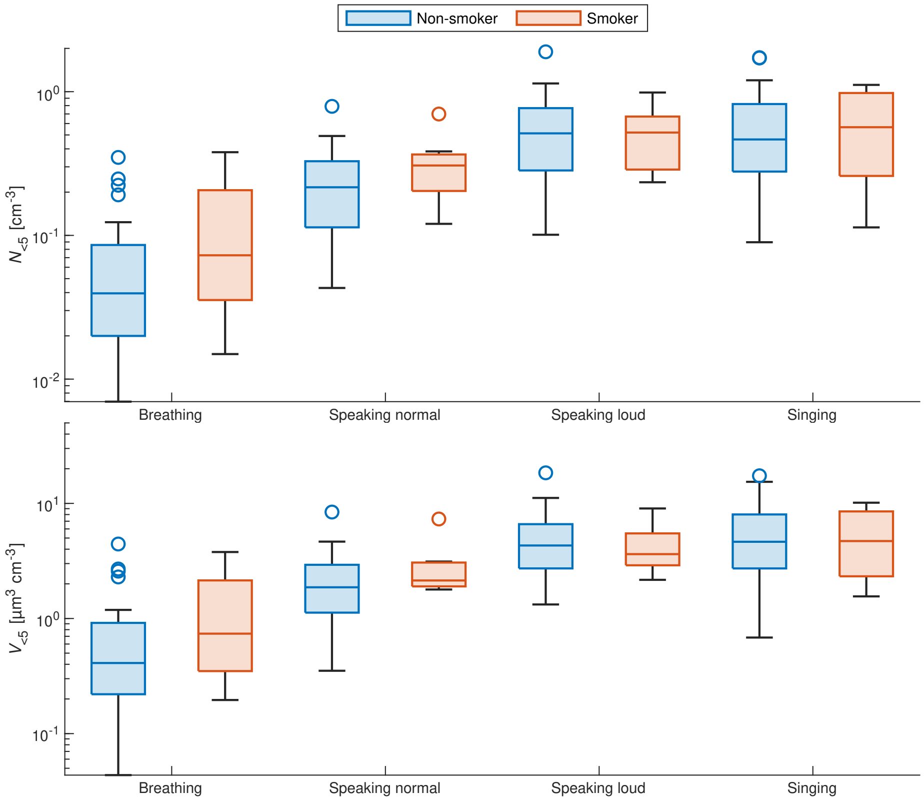


Figure S.13: Comparison between PM5 exhaled particle number and PM5 exhaled volume concentration for the standard activities between smokers and non-smokers. Compared are the smokers with all non-smokers in the test population of the same 5-year age interval.

age median of the “inactive” group is 29 years compared to 20.5 years for the “active” group. The p values differ only slightly between PM5 and PM10 used for the t-test.

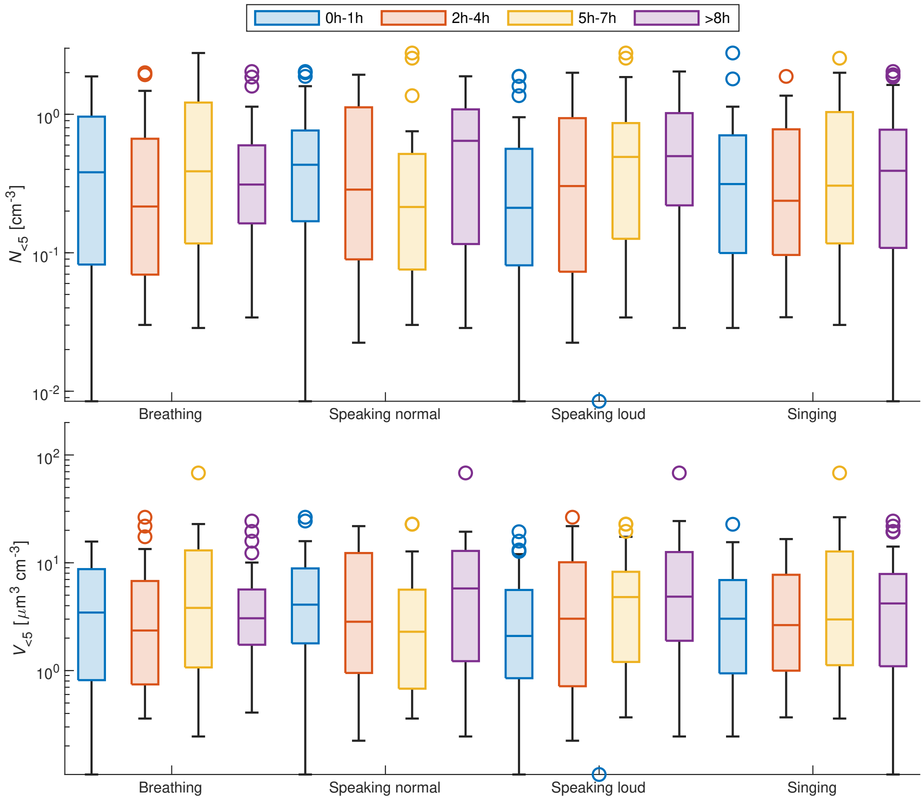


Figure S.14: Total PM5 exhaled particle number and exhaled volume concentrations in the four standard activities for subjects with different exercise habits. The times given in the legend are average hours of physical exercise per week reported by the subjects.

##### S.2.6 The correlation between Body Mass Index (BMI) and age

A naive look at our data also shows a correlation between total PM5 particle count and BMI of the subjects. However, BMI of subjects is not evenly distributed among age groups. Figure S.15 shows the total PM5 exhaled particles as a function of age similar to Figure S.12. In Figure S.15, the colour of the markers, each showing the mean PM5 particle number per activity of one subject each, indicate the BMI of the subject. Again, we can see that different BMIs span over the whole particle count range in each age group. Even though one might see a trend in the correlation between BMI and total exhaled particle number concentration for different particle sizes, by looking at the individual size-bin scatter plots ($N_i$(BMI), where $N_i$ is particle number concentration in bin $i$) we cannot confirm that the correlations actually exist and are not caused by outliers in the heavily scattered data.

When examining the particle emission for breathing as an example and selecting only subjects of 16 years or older, we see that in each subset ($n \approx 15$) defined by a BMI range (BMI between 19.3 and 22.1, BMI between 22.1 and 23.6, BMI between 23.6 and 26, and BMI between 26 and 30) there is a strong correlation between $N_{<5}$ and age for all groups and the same for $N_{\mathrm{PM10}}$. This further confirms the age-dependency of EPSD independent of BMI.

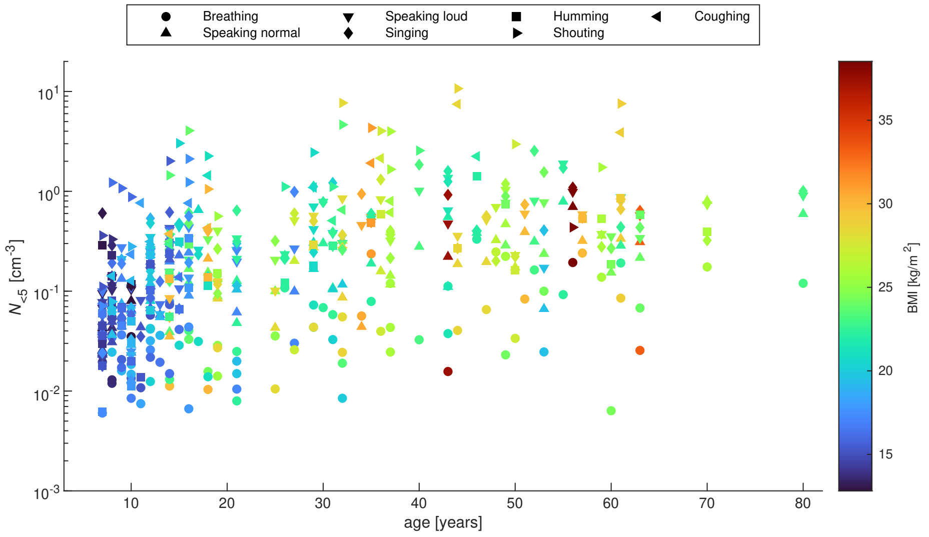


Figure S.15: Mean exhaled PM5 particle number concentration $N_{<5}$ per subject and activity as a function of age. The colour of the individual markers indicate BMI of the subject.

##### S.2.7 Temporal variability of respiratory particle emission

The distribution of $N_{<5}$ and $V_{<5}$ over a time period of 60 to 240 s for samples of 1 s (sampling interval $\ll$ duration of respiratory event) was found to be log-normal for all activities done by the same subject. As the geometric mean $\mu_g$ could be determined from a log-normal fit for each activity examined, the values for the different $\mu_g$ were compared to the mean $N_{<5}$ and $V_{<5}$ which were usually obtained from samples of 60 s duration. The ratio between $\mu$ and the average $N_{<5}$ and $V_{<5}$ was very close to 1, which shows that averaging over a time scale long enough in comparison to the duration of a single event (one breathing cycle or speaking one syllable or singing one note) actually yields the emission median.

Our 60 s samples are therefore a good representation of the average respiratory particle emission during all activities we have studied. On the other hand, we saw that the 95th percentile of $N_{<5}$ is usually a factor of 2 above the median for the phonetic activities (speaking, singing, humming, shouting, coughing), and about a factor of 5 above the median for breathing. In terms of $V_{<5}$, the 95th percentile is about a factor of 5 higher than the median for the phonetic activities, and about a factor of 10 higher than the median for breathing. In case of deep breathing (described in Table S.2), the distribution of $N_{<5}$ and $V_{<5}$ is similar to normal breathing but with much higher values (see Fig. S.16).

##### S.2.8 Total number and volume statistics and the question of super-emission

From all experiments, we investigated the distribution of total PM5 number concentration $N_{<5}$ and total PM5 volume concentration $V_{<5}$ among the subjects. If a subject did multiple experiments of the same kind, the median of $N_{<5}$ and $V_{<5}$ from all experiments of this subject was taken. Fig. S.17 and Fig. S.18 show the cumulative distribution functions of $N_{<5}$ and $V_{<5}$ respectively from all subjects and experiments with the isolation shield. For both the entire cohort and for each age group as described below, we found that $N_{<5}$ and $V_{<5}$ could follow both Gaussian or log-normal distributions as the quality of fit measured by $r^2$ was above 0.9 for both distribution types. However, a log-normal distribution is in all cases the better representation of the data compared to a Gaussian, in particular for the tails. We calculated the geometric mean $\mu_g$ and the geometric standard deviation $\sigma_g$ for $N_{<5}$ and $V_{<5}$ for each kind of experiment and computed a log-normal distribution with the same $\mu_g$ and $\sigma_g$. The log-normal distribution was then compared with the measured distribution by performing a two-sided Kolmogorov-Smirnov test. In all cases, the null hypothesis could not be rejected at 95% significance level, which means that the observed distributions can be approximated well by log-normal distributions. In addition, we fitted log-normal distributions to the data and got good fits ($r^2 \geq 0.91$) with very little deviation in $\mu_g$ and $\sigma_g$ from the values computed directly from the data. An overview of the results from the fits is shown in Table S.7. As there is an age dependence in $N_{<5}$, we needed to examine the distribution of emitted $N$ among the subjects in subsets defined by age. The strongest increase of $N_{<5}$ as a function of age was seen for subjects below 20 years (see Fig. S.12), so we decided to subdivide the data pool in 5 year age intervals from 5 to 19 years, and in 10 year age intervals for the adults of age 20 years or more. The last age group in Table S.7 contains all subjects between 50 and 80 years as the total number of subjects in this age category was too small to justify smaller age subgroups. As already discussed in the subsection on subject variability in the main paper, we see an increase in the geometric mean of $N_{<5}$ for increasing age, which is strongest for the louder phonetic activities (loud speaking and singing). The numbers of low- and super-emitters (see definition above Subsection S.2.1) in the different activities, as well as the number of low- and super-emitters in all categories, are presented in Table S.8.

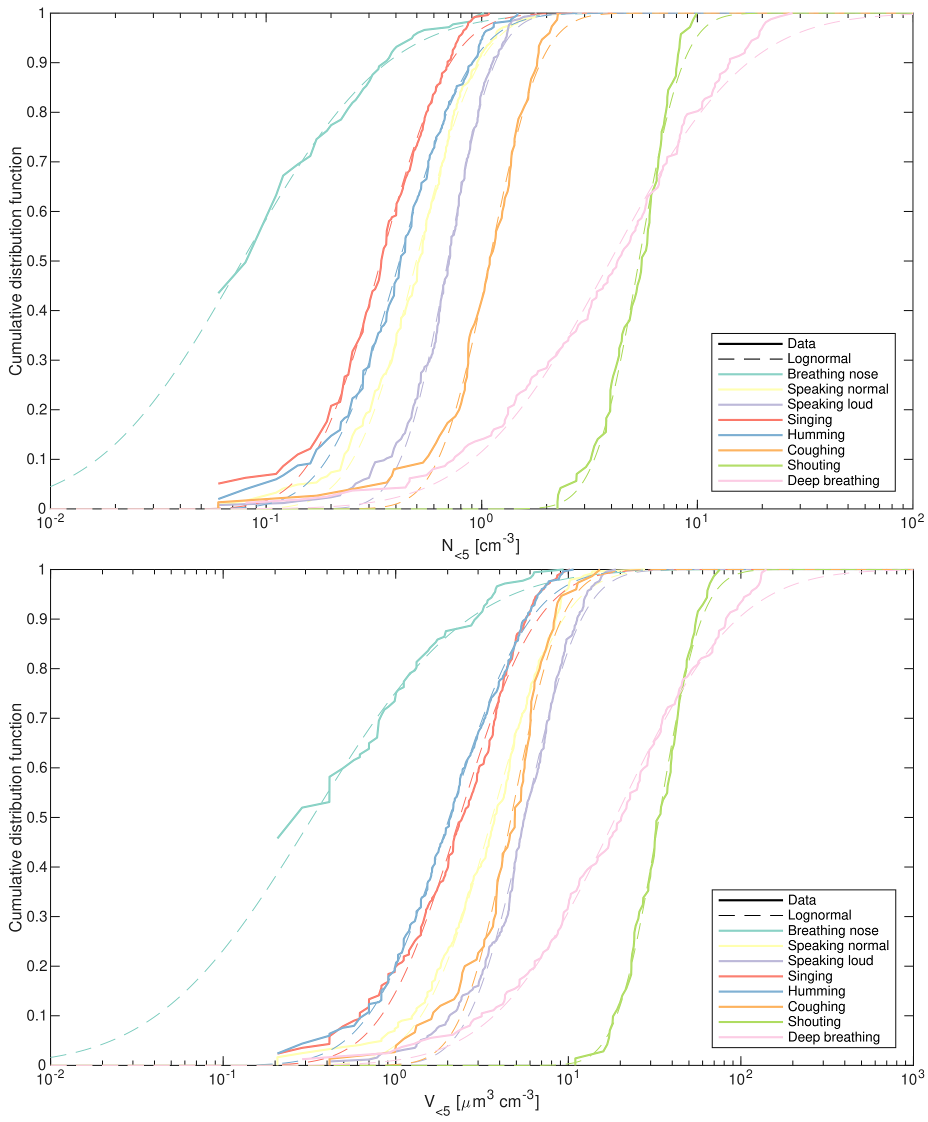


Figure S.16: Cumulative distribution function of measured $N_{<5}$ (top) and $V_{<5}$ (bottom) on one subject with 1 s sampling. The data (solid lines) and log-normal fits (dashed lines) are shown for breathing through the nose (green), speaking normal (yellow), speaking loud (purple), singing (red), humming (blue), coughing (orange), shouting (light green), and deep breathing (pink).

| age | breathing | | speaking n. | | speaking l. | | singing | |
|---|---|---|---|---|---|---|---|---|
| | $\mu_g$ | $\sigma_g$ | $\mu_g$ | $\sigma_g$ | $\mu_g$ | $\sigma_g$ | $\mu_g$ | $\sigma_g$ |
| 5-9 | 0.024 | 1.781 | 0.041 | 1.654 | 0.072 | 1.859 | 0.068 | 2.675 |
| 10-14 | 0.020 | 2.128 | 0.077 | 2.324 | 0.120 | 2.187 | 0.129 | 2.769 |
| 15-19 | 0.022 | 1.847 | 0.137 | 1.594 | 0.220 | 1.938 | 0.220 | 2.383 |
| 20-29 | 0.027 | 2.398 | 0.131 | 1.704 | 0.314 | 1.756 | 0.391 | 2.184 |
| 30-39 | 0.043 | 1.966 | 0.140 | 1.916 | 0.401 | 1.486 | 0.428 | 1.803 |
| 40-49 | 0.050 | 2.921 | 0.233 | 1.611 | 0.560 | 1.588 | 0.605 | 2.171 |
| 50+ | 0.091 | 2.879 | 0.313 | 1.964 | 0.571 | 1.938 | 0.578 | 1.872 |

Table S.7: Geometric mean $\mu_g$ in cm$^{-3}$ and geometric standard deviation $\sigma_g$ of $N_{<5}$ for each age group in all four activities obtained from a log-normal fit to the data.

| age | $n_{sub}$ | breath. | | speak. norm. | | speak. loud | | singing | | in all act. | |
|---|---|---|---|---|---|---|---|---|---|---|---|
| | | low | high | low | high | low | high | low | high | low | high |
| 5-9 | 18 | 3 | 2 | 5 | 4 | 3 | 2 | 3 | 3 | 0 | 0 |
| 10-14 | 25 | 3 | 4 | 4 | 6 | 2 | 3 | 4 | 4 | 0 | 1 |
| 15-19 | 14 | 2 | 3 | 2 | 1 | 4 | 3 | 3 | 2 | 0 | 0 |
| 20-29 | 20 | 4 | 4 | 3 | 3 | 1 | 3 | 2 | 4 | 0 | 1 |
| 30-39 | 15 | 2 | 2 | 1 | 3 | 2 | 3 | 3 | 3 | 0 | 0 |
| 40-49 | 13 | 2 | 3 | 2 | 2 | 1 | 3 | 3 | 2 | 1 | 0 |
| 50+ | 21 | 4 | 2 | 2 | 4 | 3 | 3 | 4 | 3 | 0 | 0 |

Table S.8: Number of low and super-emitters in the different age groups. $n_{sub}$ denoted the number of subjects for this analysis per age group. The last two columns show how many subjects were low- or super-emitters in all four considered activities.

In conclusion, we investigated the relationship between extraordinarily high and low emission and activity for seven age groups and we found that on average only about 0.8 - 1.6 % of the subjects tend to be either "global super-emitters" or "global low-emitters", which means that they are well below or above the median emission in all four activities examined. The percentage of super-emitters in one activity is between 16.4 % (breathing) and 18.2 % (speaking normal), the percentage of low-emitters is between 13.9 % (speaking loud) and 18.3 % (singing). With an average of 17.2 % for super-emitters and 15.8 % for low-emitters, both extremes are nearly equally distributed within the population. What we also found is that 11/20 (55 %) of the low-emitters in breathing are also low-emitters in at least one phonetic activity, and 10/20 (50 %) of the super-emitters in breathing are also super-emitters in at least one phonetic activity. The abundance within the examined population of the super-emitters in breathing plus one phonetic activity is 8.0 % and the abundance of low-emitters in breathing plus one phonetic activity is 8.4 %.

The most extreme low-emitter (i.e. the minimum from all age groups and activities) had a value of $N_{<5}$ which was 2.7 geometric standard deviations below the (global) geometric mean of $N_{<5}$. The most extreme super-emitter was 3.1 geometric standard deviations above the geometric mean of $N_{<5}$. On average, the most extreme low-emitters and super-emitters per age group were 1.69 and 1.86 geometric standard deviations away from the geometric mean of $N_{<5}$. However, the data for the low-emitters should be interpreted with care as very low emission might have other causes like a mask not perfectly sealing. When doing the naive approach, which is taking all breathing, speaking and singing data independently of the age of the subject, the most extreme low-emitter is 3 geometric standard deviations below the geometric mean, and the most extreme super-emitter is 3 standard deviations above the geometric mean.

##### S.2.9 Additional test measurements

In the following, the results of different-test experiments are analysed to investigate different factors that potentially influence the EPSD measurements with spectrometers. Each test experiment was conducted with 1-3 subjects. For the analysis, OPS data was used. The results are therefore in the size range of 0.3 µm to 10 µm dry diameters (1.36 µm to 45.5 µm exhaled diameter), a smaller particle diameter range is shown when no particles were counted in the larger bins. In most-test experiments, two OPS were used together (similar to S.1**d** but with a second OPS instead of SMPS) resulting in a sample flow rate of $2\,\mathrm{l\,min^{-1}}$. The shown EPSD are each the mean of both OPSs, whereas the time series data is only from one single OPS. In these experiments sampling frequency was $1\,\mathrm{s^{-1}}$. In sec. S.2.9.5, the setup Fig. S.1**d** was used, only the OPS data is analysed and shown. In section S.2.9.2-S.2.9.7, the sampling was done with the new isolation shield. The advantage of these test experiments is that the comparisons are made for activities/setups on the same subjects each, therefore only within-subject variability has to be considered but not the higher between-subject variability.

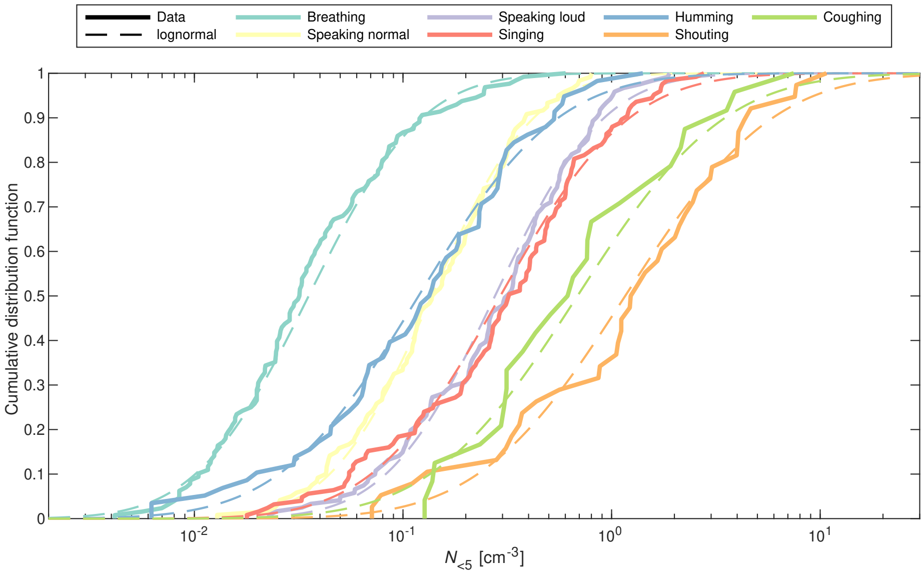


Figure S.17: Cumulative distribution function of $N_{<5}$ from all experiments done in the MPI-DS cleanroom with the isolation mask. The solid lines represent the measured data, the dashed lines represent the log-normal distributions.

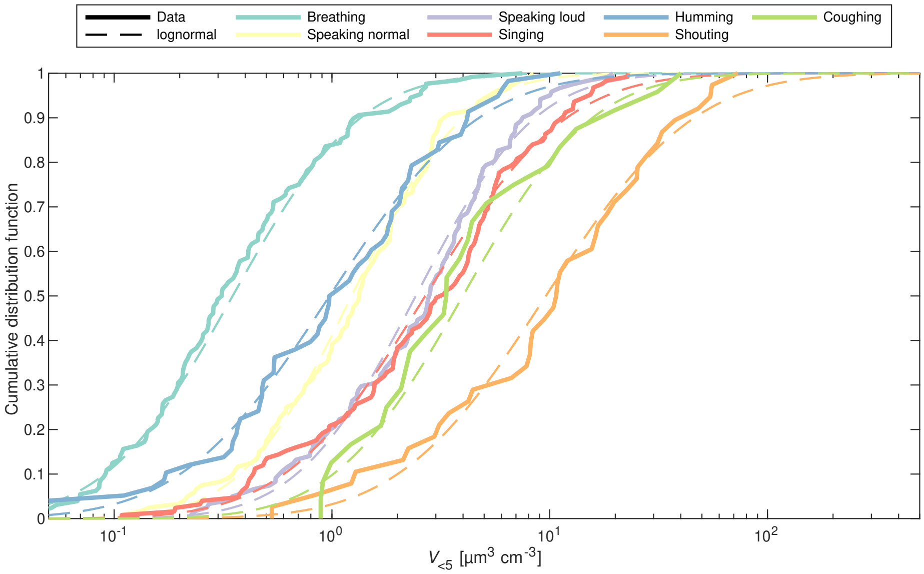


Figure S.18: As in Fig. S.18 for $V_{<5}$.

###### S.2.9.1 Effect of isolation shield model

Figure S.19 shows a comparison between the two different isolation shields that were used (see S.1). The test measurement was performed on two different subjects, the results of each subject are shown individually. For better visibility, no error bars showing the uncertainties related to counting statistics are shown. Generally, particle counts are higher and therefore the measurement uncertainties are smaller for the male super-emitter over the female low-emitter and for smaller over larger particles. The data suggest that fewer particles are lost in the new isolation shield for a male super-emitter especially in the vocal activities which could be explained by the less complex shape of the exhale cavities (see Fig. S.1 **a** and **b**). However, due to high variability in the data (cf. sec. S.2.2) it is unclear how significant this deviation is, we also see no trend – and for large particles even the opposite – for the female low-emitter, where overall fewer particles were exhaled.

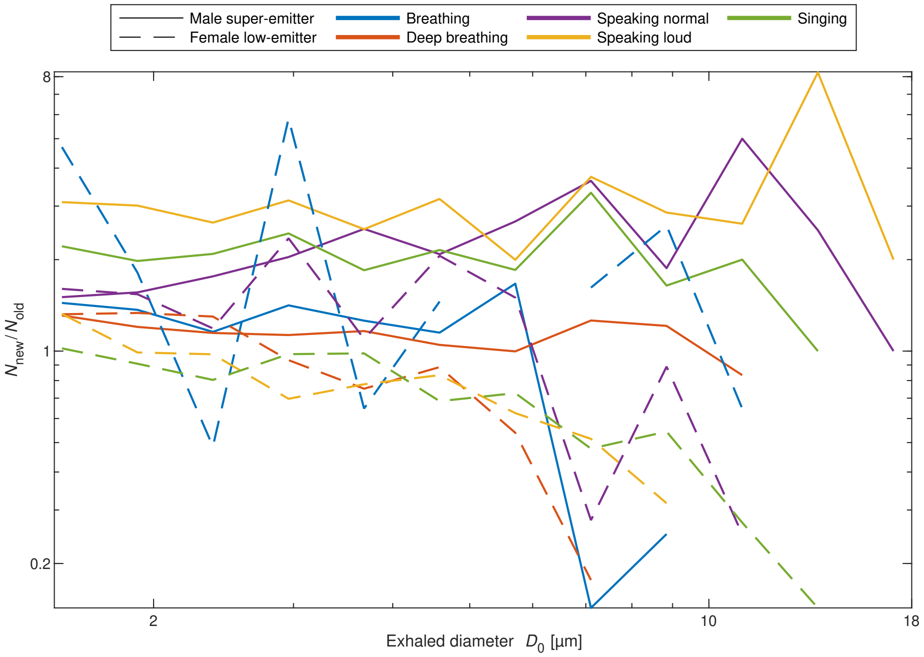


Figure S.19: Comparison of EPSD sampled with two different models of isolation shields. Results for two individual subjects performing the standard activities and deep breathing are shown.

###### S.2.9.2 Effect of buffer volume size

On the same two subjects, another test experiment was conducted to investigate the influence of the buffer volume size. In the original setup, the buffer volume on the exhale was $\sim 30$ ml with the new isolation shield plus 4 ml in the exhalation filter adaptor. The buffer volume of the old isolation shield adaptor is >80 ml (buffer volume inside the isolation shield excluded). The exhale buffer volume was increased with a tube with a diameter of 5/8"=1.59 cm and 1.06 m or 2.01 m length (i.e. by volumes of 0.21 l and 0.40 l respectively). When these additional tubes were used, the exhalation filter was removed to avoid increasing the exhalation pressure too much. Figure S.20**a** shows the results for different activities of the experiments with additional buffer volume normalised with particle counts of the original setup with small buffer volume and filter. It is evident that for activities where the exhalation time significantly exceeds the inhalation time (i.e. reading, singing, shouting) a larger buffer volume makes no difference. For these activities, it can therefore be concluded that already in the original setup only the exhalation concentration was measured and no mixing with cleanroom air occurred despite the small buffer volume.

For breathing and deep breathing, however, the buffer volume makes a difference. Within-subject variability has to be considered here as well, however, the minimal scatter in the other activities, as well as the following discussion about the time series, indicate an actual effect of a larger buffer volume. The measured particle concentrations are higher by up to a factor of 4 with additional tubes (for the female low-emitter in both cases for the male super-emitter only with an additional volume of 0.4 l). This can be explained by the relatively long inhalation periods in breathing activities. During inhalation, the sampling flow empties the buffer volume (assuming linear flow, 34 ml exhaled air would be completely emptied within 1 s with the flow rate of 2 l min$^{-1}$; within 1.2 s with setup Fig. S.1**d** and 0.3 s with setup Fig. S.1**e**). If the pressure difference produced by the sampling flow rate becomes large enough and no exhalation follows, cleanroom air (no particles) is sucked through the filter into the buffer volume and is sampled (particle count drops to zero). In a frequency-controlled measurement, we found that each exhalation period typically ends shortly after the observed peaks in Fig. S.20**b** (in between peak pairs when measured with buffer tube, particle delay due to tubes are taken into account, i.e. peak particle production is not during the end of exhalation), this observation can also be applied to these non-controlled measurements. After exhalation stops, with the larger buffer volume, exhaled air that is stored in the long tube is sampled as opposed to clean air with no buffer volume. In the deep breathing case, where the inhalation period is very long, this leads to a typical "double peak" (cf. Fig. S.20**b**, middle and bottom). This is caused by the exhaled air not mixing well in the tube and therefore the sampling "looks backwards" in time (air exhaled right before exhalation stops is in the beginning of the tube and therefore gets sampled first, air with highest particle count was exhaled earlier and is, therefore, further down in the tube and gets sampled later) and reproduces the original peak. This effect is more pronounced for the longer tube since mixing with clean air occurs less and later. The same effect likely happens in normal breathing too but cannot be observed due to limited time resolution and lower emitted particle number. Since this effect was only tested on two subjects and considering the high within-subject variability (sec. S.2.2) the stated

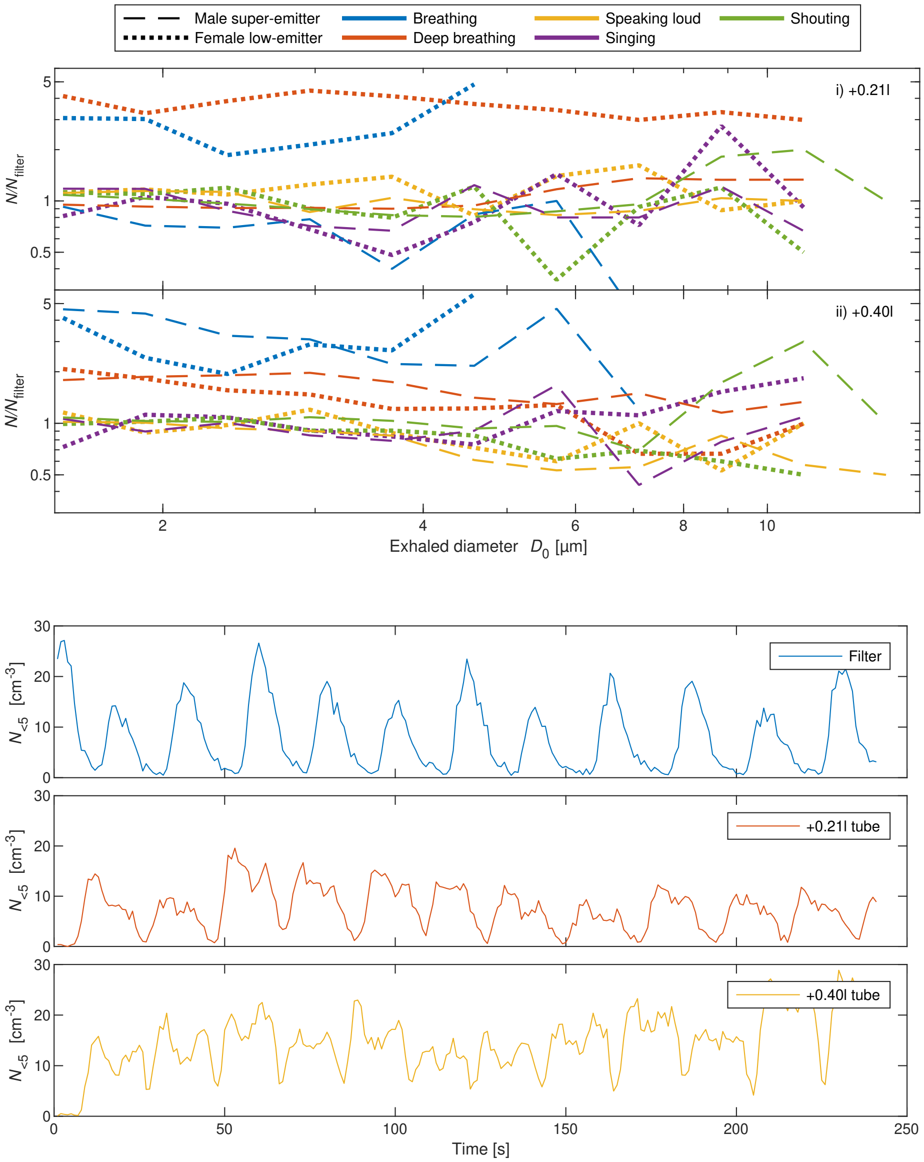


Figure S.20: Effect of an additional buffer volume on the exhale. **a** EPSD with buffer volume are shown normalised with particles counts in the original setup (just the filter on the exhale). The results are shown for the two subjects individually. **b** Total PM5 particle count time series for deep breathing of a male super-emitter (high emitter, therefore clear signal) for different buffer volumes. The lowest minima occur right before exhalation starts. With additional buffer volume on the exhale another smaller peak occurs after the end of exhalation.

factor of up to 4 has to be treated cautiously. Assuming inhalation and exhalation periods have approximately the same length, the effect of underestimating should be by a factor of $\leq 2$. The measured exhaled particle concentrations in breathing activities can still be interpreted as average over inhalation and exhalation. It has to be noted, that the only way to effectively measure the particle concentration in the exhale during breathing activities (where inhalation time $\approx$ exhalation time) would be to only sample during exhalation. With deep breathing sampled with a high sampling frequency and subjects exhaling high particle concentrations (e.g. as shown in S.20**b**), the exhalation times could also be later defined in the analysis and the other samples could be excluded. However, most subjects had significantly lower particle concentrations and higher breathing frequencies, so this detection of exhalation periods was not possible.

###### S.2.9.3 Effect of breathing frequency

We measured the exhaled particle concentration for different breathing frequencies. The exhalation to inhalation duration ratio was kept constant with the help of a breath visualisation video (section S.1.2). The video was played at 4 different speeds, resulting in 4 different breathing frequencies. There was no pausing in between inhalation and exhalation or exhalation and inhalation and subjects tried to perform both full inhalation and full exhalation in all frequencies. Figure S.21 shows the particle counts of lower breathing frequencies normalised with the particle counts of the highest frequency denoted $N_f$. The measurements were performed with and without buffer volume and on 1-3 subjects. Measurements with/without additional buffer volume were normalised with measurements with/without buffer volume respectively. This way, the effect discussed in S.2.1.2 should not make a difference. We can see that the individual results are scattered around 1 and no clear trend can be observed. The scattering can be explained by the within-subject variability.Therefore we conclude, that breathing frequency does not affect respiratory particle production.

###### S.2.9.4 Effect of breathing patterns

Figure S.22 shows the time series of the counted particles for two different breathing patterns for two subjects, a male super-emitter and a female low-emitter. We compare full inhalation and exhalation with a 10s pause in between to no pause in between. The measurements were performed with the long tube as an additional buffer volume of 0.4 l. Since breathing frequency was not monitored here and in the case without pause there is a higher exhalation time relative to the whole sampling time, the bias discussed in section S.2.1.2 has to be kept in mind when comparing the particle size distributions (Fig. S.22, top). However, comparing the individual peak heights of the time series of exhaled PM5 particles (Fig. S.22, middle and bottom) confirm the trend that can be seen in the particle size distributions that pausing between full inhalation and exhalation significantly decreases the exhaled particle count. This effect has

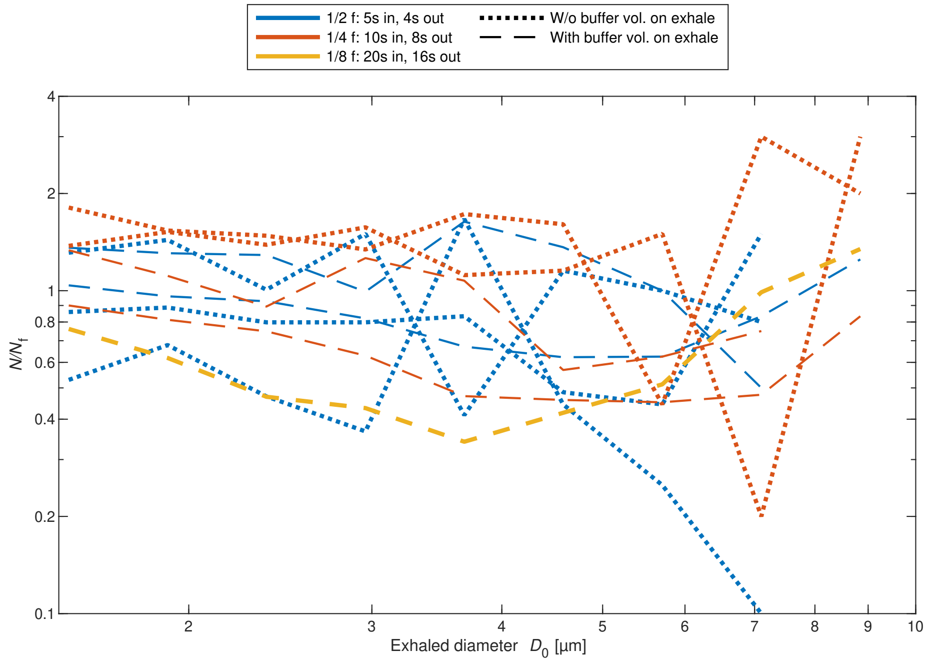


Figure S.21: EPSD for different breathing frequencies normalised with EPSD for the fastest frequency used as a reference. Inhalation to exhalation ratio is kept constant. Results for different subjects are shown individually.

been previously studied in more detail in several previous investigations [1].

Another breathing pattern that was investigated was shallow breathing with an exhalation every second ($f = 1\,\mathrm{s}^{-1}$). In Figure S.23 it is compared to controlled deep breathing (5 s inhalation, 4 s exhalation) with the help of the breath visualisation video. For two of the three subjects particle emission is significantly decreased, whereas for one it increases.

###### S.2.9.5 Effect of external acoustic excitation

In one series of experiments, we investigated the possible impact of external acoustic excitation of the chest region of one subject. The sound that was used was a 110 Hz sine wave with 80 dB(A) at 1 m distance from the loudspeaker. The loudspeaker was placed 0.1 m from the chest of the subject. Three activities were examined, which were normal breathing through the nose, singing "Happy Birthday" with an open mouth and humming "Happy Birthday" with a closed mouth. Fig. S.24 shows the measured EPSD for each activity. We found minute differences between the EPSDs for the same activity with and without the external sound, which are all within the uncertainties from counting statistics. As the frequency that was examined was close to the subject's chest resonance (the central pitch of the subject's speaking voice is between the note Ab2 and Bb2), it is highly unlikely that there is a difference for any other frequency at the same sound pressure.

###### S.2.9.6 Effect of vocalisation

To investigate the effect of vocalisation and therefore the role of the larynx in particle emission we performed a deep breathing experiment on 2 subjects with and without vocalisation. The silent case is a deep breathing activity synced to a breathing visualisation video, whereas the loud case is with a vocalisation of "aah" during the whole exhalation of the same breathing pattern (cf. Table S.2). Figure S.25 shows the ratio of the exhaled particles during the activity with and without vocalisation for both subjects individually and the mean. The vocalisation on the exhale does not show a significant difference, in particular for $D_0$ <3 µm where strong statistical convergence could be achieved. Since breathing frequency was kept constant over the silent and loud breathing activity and in both cases the exhale was a full exhale.

###### S.2.9.7 Size-dependent arrival times

From a standardised breathing pattern where the subject breathing was in sync with the breathing visualisation video (section S.1.2), we had a well-known periodicity and could investigate a possible phase-shift between different particle sizes. The OPS device was set to measure one sample each second (the minimum sampling duration available in the instrument). Each exhale pattern start was defined as the time where the total particle concentration was at zero before it rapidly increased. The duration of the exhale period was 15 s in this experiment.

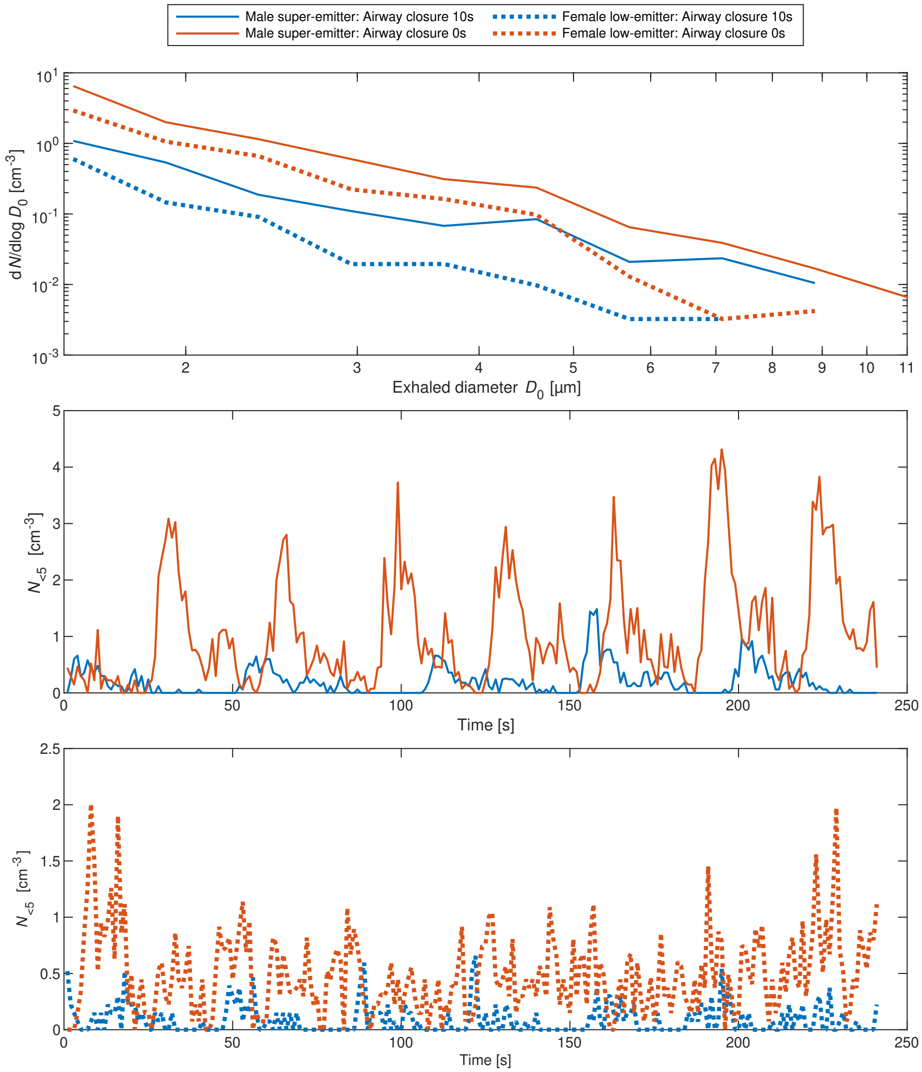


Figure S.22: Total PM5 particle count time series for full in- and exhalation breathing pattern with and without pausing i.e. holding breath in between. The particle size distribution and time series for the two tested subjects are shown individually. Pausing reduces the exhaled particle concentration significantly.

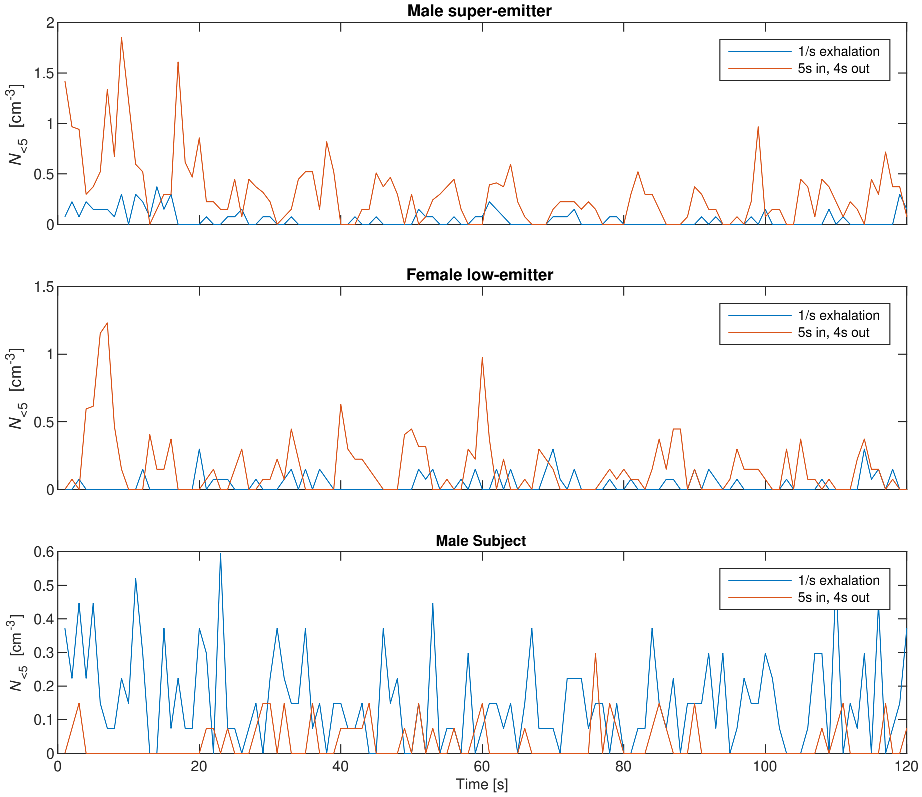


Figure S.23: Comparison of total PM5 particle count time series between a shallow breathing pattern with exhalation every second and a full in- and exhalation breathing pattern (5s in, 4s out). The time series for the three tested subjects are shown individually.

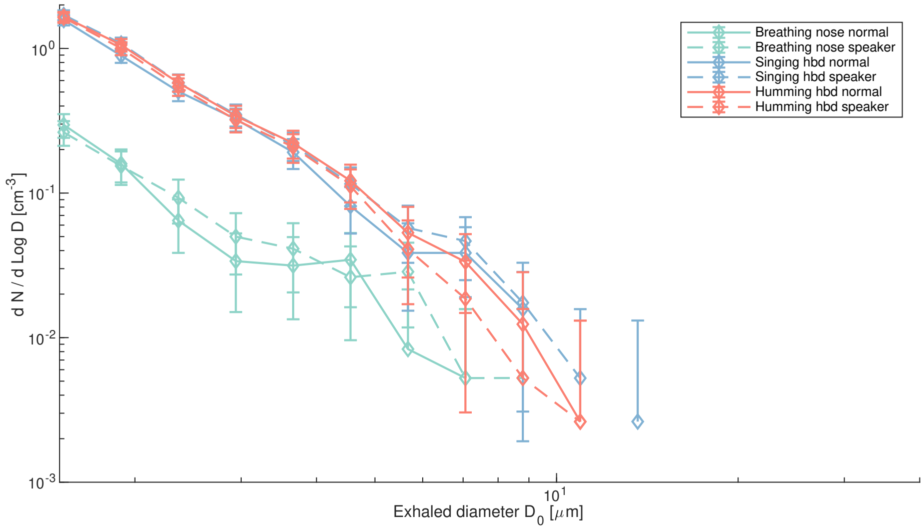


Figure S.24: EPSD for experiments with and without external acoustic excitation. The experiments represented with dashed lines (speaker) are with external sound, the others with the solid lines were without external sound. Shown are the EPSDs for breathing (green), singing "Happy Birthday" (blue), and humming "Happy Birthday" (red). Errorbars represent counting statistics for each experiment (Poisson distribution assumed).

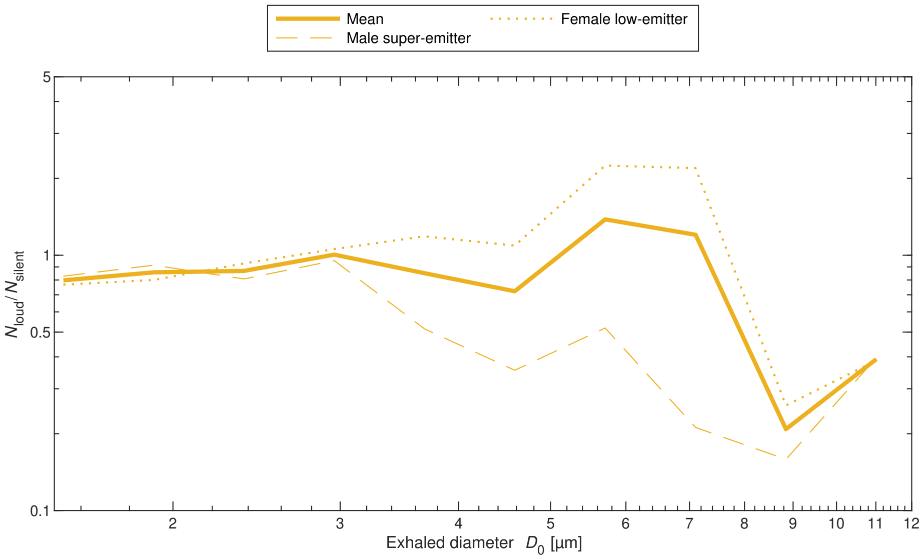


Figure S.25: Effect of vocalisation during deep breathing on EPSD. Subjects vocalised "aah" during exhalation. Solid lines show the mean over the two subjects. Results for individual subjects are shown in dotted lines as a measure of uncertainty. Details of activities are presented in Table S.2.

When comparing particles smaller than 2.1 µm, between 2.1 and 5.1 µm and larger than 5.1 µm in terms of time of arrival, we see that the maximum in the PDF show a trend towards the larger particles arriving before the smaller ones, and the intermediate size reaches the median in the CDF about 3 s before the large size (see Fig. S.26 and S.27). The time shift in the median is approximately 1 s between the small and large particles while the maxima in the PDF are up to 6 s apart from each other (the largest particles at 5 s, the intermediate size at 6 s, the smallest at 11 s). This example is taken from one subject. Another subject did the same breathing pattern, with the same qualitative result but lower particle number concentration. The increase in particle concentration after the stop of exhalation is due to the buffer volume that was attached to the isolation shield. We need to emphasise that the observed phase shift cannot be explained by the flow dynamics caused by the internal structure of the mask and the tubing, but as the maxima for the large and intermediate size are only one sample apart from each other, we are already at the maximum resolution of the data. It also needs to be mentioned that counting statistics are much lower for exhaled diameters of larger than 5 µm compared to PM5. Particle by particle data from a PSS with much higher temporal resolution are needed to investigate this phase shift further.

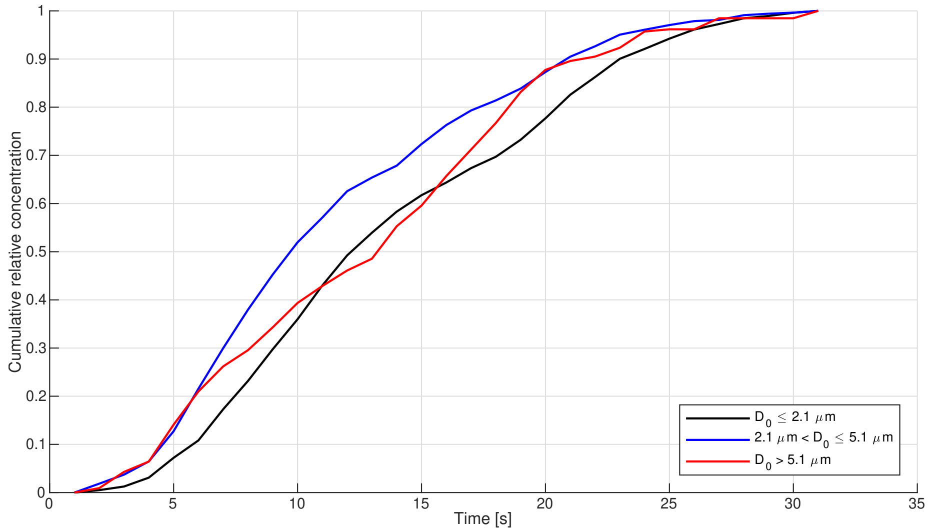


Figure S.26: Mean cumulative distribution function of particle concentration for small (black), intermediate (blue) and larger size (red) as a function of time for periodic slow breathing.

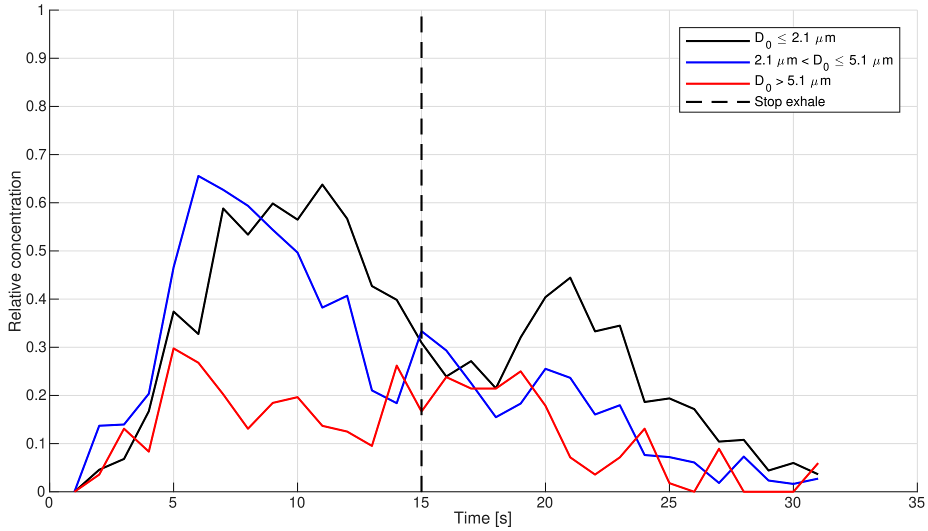


Figure S.27: Mean probability density function of particle concentration for small (black), intermediate (blue) and larger size (red) as a function of time for periodic slow breathing.

### References

[1] B. Bake, P. Larsson, G. Ljungkvist, E. Ljungström, and A-C Olin. Exhaled particles and small airways. *Respiratory Research*, 20:8, 12 2019.

[2] Bastian Bechtold, Patrick Fletcher, seamusholden, and Srinivas Gorur-Shandilya. bastibe/violinplot-matlab: A good starting point, February 2021.

[3] Jitendra K Gupta, Chao-Hsin Lin, and Qingyan Chen. Characterizing exhaled airflow from breathing and talking. *Indoor air*, 20(1):31–39, 2010.

[4] Oliver Schlenczek. *Airborne and ground-based holographic measurement of hydrometeors in liquid-phase, mixed-phase and ice clouds.* PhD thesis, Particle Chemistry, Max Planck Institute for Chemistry, Max Planck Society, 2018.
